# Prevalence of mental disorders in people with intellectual disabilities across the lifespan: an umbrella review

**DOI:** 10.1101/2025.09.21.25336181

**Authors:** Jacopo Santambrogio, Giovanni Boido, Mattia Marchetti, Sonya Rudra, Filippo Besana, Emma Francia, Sergio Terrevazzi, Davide Papola, Corrado Barbui, Marco O. Bertelli, Massimo Clerici, Armando D’Agostino, Louise Martson, Angela Hassiotis

## Abstract

**Background:** People with intellectual disabilities (ID) experience higher rates of mental disorders, contributing to restrictive practices and premature mortality. Prevalence data are essential to inform support and policy decisions.

**Aims:** To examine the prevalence of mental disorders in people with ID across the lifespan.

**Methods:** We systematically searched six databases and conducted a manual search up to December 15^th^, 2024, to identify systematic reviews on the prevalence of mental disorders in people with ID of any aetiology with or without other neurodevelopmental conditions. We summarised prevalence data from systematic reviews with meta-analyses and from systematic reviews with a narrative synthesis of results, including subgroup analyses (ID level, sex, age, autism), where possible. We compared these results with published prevalence rates of mental disorders in people without ID. PROSPERO: CRD42024610611.

**Results:** We included twenty-six systematic reviews: seven with meta-analyses, and nineteen with subgroup or narrative synthesis. Compared to the general population, people with ID showed higher prevalence of schizophrenia (3.55%-4.8%), anxiety (5.4%-5.5%), and obsessive-compulsive (2.4%) disorders, while mood disorders (6%-7%), personality and post-traumatic stress disorders were less frequent, though data were limited. Study quality was moderate to critically low (AMSTAR-2). An emerging picture of mental disorders is evident in syndromic ID, with high anxiety rates in fragile X, Williams, and 22q11.2 deletion syndromes, and in co-occurring autism. This umbrella review revealed gaps in research on other mental disorders (i.e., dementia, bipolar, substance use, and eating disorders) and a lack of prevalence data stratified by age, sex, and ID level. This is compounded by the absence of dependable diagnostic criteria suitable for people with ID.

**Conclusions:** This umbrella review confirms the high prevalence of mental disorders among people with intellectual disabilities, and highlights limited evidence for several conditions and population subgroups. Standardised, high-quality epidemiological research is needed to guide clinical practice and policy.

## BACKGROUND

Intellectual disabilities (ID) are lifelong conditions accounting for significant limitations in intellectual functioning and adaptive behaviour across cognitive, social, and practical domains with onset during the developmental period, typically in preschool (1, 2). The estimated global prevalence of “idiopathic intellectual developmental disability” is around 2% (3) and the incidence around 1.8% (4).

People with ID experience significant health inequalities. These are often the result of barriers to specialised healthcare, diagnostic overshadowing, stigma, and limited access to education and employment (5). Primary studies of people with ID report significant rates of mental comorbidities ranging from 30% to 50% (including behaviours that challenge) (4); with Cooper et al. (6) identifying a point prevalence of mental disorders of 40.9% (clinical diagnoses), 35.2% (using diagnostic criteria for learning disability-DC-LD), 16.6% (ICD-10-DCR) and 15.7% (DSM-IV-TR) in a population study. Mazza et al. (7) highlighted a prevalence rate of 33.6% in clinical and community samples of adults with ID, whilst Einfeld et al. (8) reported 30%-41% for mental disorders in children and adolescents with ID, compared to 8%-18% in those without. Similarly, Emerson et al., found a prevalence of 36% in children with ID versus 8% in those without (9).

Determining the prevalence of specific mental disorders in people with ID with or without others neurodevelopmental disorders (e.g., autism spectrum disorder, ASD) is particularly challenging, due to historical variations in diagnostic systems, difficulties in finding and defining representative samples (10, 11), the limited availability of tailored diagnostic criteria for people with ID, and the complex interface between behaviours and mental disorders (12).

While people with ID are known to experience the full spectrum of mental disorders, synthesising existing evidence is essential to identify gaps, guide research priorities, improve clinical awareness, and facilitate access to tailored interventions for this underserved group.

To our knowledge, although two umbrella reviews have explored both mental (13) and physical (14) comorbidity in autistic people, similar endeavours are lacking in people with ID despite a growing body of research on this topic.

The research questions were:

1. What is the prevalence of specific mental disorders in people with ID without comorbid neurodevelopmental disorders?
2. What is the prevalence of specific mental disorders in people with ID with other neurodevelopmental disorders (e.g., ASD)?
3. What are the gaps in the literature on the prevalence of comorbid mental disorders in people with ID with or without other neurodevelopmental disorders?

Where available, we have compared prevalence rates in people with ID with those in the general population.

## METHODS

We conducted this umbrella review according to the Preferred Reporting Items for Systematic Reviews and Meta-Analyses (PRISMA) guidelines (15) (Supplementary Material, Table 9-10), and the protocol was registered with the International Prospective Register of Systematic Reviews (PROSPERO, reference: CRD42021230023) (16).

### Literature sources

J.S. developed a systematic search strategy assisted by G.B. and by the bibliographic services of the University of Milano-Bicocca. “Developmental Disabilities”, “Intellectual Disability”, “Persons with Mental Disabilities”, “Learning Disabilities”, “Specific Learning Disorder”, and “Comorbidity” were used as MeSH Terms and adapted through six databases: the Cochrane Library, PubMed, PsycINFO (via ProQuest interface), Embase, CINAHL and Epistemonikos. Filters for Systematic Reviews, or Meta-analysis were applied when appropriate and available. G.B. and M.M. searched the grey literature on the following web platforms: WPA (World Psychiatric Association), EPA (European Psychiatric Association), NADD (National Association for the Dually Diagnosed), EAMHID (European Association for Mental Health in Intellectual Disability), IASSIDD (International Association for the Scientific Study of Intellectual and Developmental Disabilities), and WHO (World Health Organization). In addition, we performed manual searches for all articles selected at full-text screening through backward and forward citation tracking, with searches conducted up to mid-December 2025. (Supplementary Material, Table 1, for further details on the search strategy).

### Eligibility Criteria

We included studies if they: 1) were a systematic review of observational studies (cross-sectional, cohort, ecological studies), 2) summarised the prevalence of different mental disorders, 3) were conducted in children and/or adults with ID of any aetiology.

Reviews that primarily investigated other populations (e.g., autistic people) were included if they presented studies in which at least 50% of the sample participants had an ID, and if they reported prevalence data for this group. We included genetic syndromes most frequently associated with a high (or near-complete) penetrance of ID, namely Down syndrome, Fragile X syndrome, Williams syndrome, 22q11.2 deletion syndrome, Prader-Willi syndrome, while excluding reviews that focused predominantly on genetic investigations or on rarer syndromic types.

Book chapters and textbooks were excluded unless they explicitly referred to systematic reviews. Theses and conference abstracts with systematic searches were excluded; however, we searched for corresponding peer-reviewed full-text publications for potential inclusion. We did not report prevalence estimates of ASD or Attention-Deficit/Hyperactivity Disorder (ADHD) in people with ID when these conditions were presented as mental disorders in people with ID without providing further specific information on the co-occurrence of other mental disorders in people with both ID and another neurodevelopmental condition, such as ASD or ADHD.

### Procedure

We undertook a literature search on six databases from inception and a manual search up to December 15, 2024. Articles retrieved from the database search were exported into EndNote 21, and after deduplication, two groups of reviewers (J.S.-E.F.-F.B.; G.B.-M.M.) independently screened titles and abstracts for full-text inclusion and conducted the final study selection. G.B. conducted the manual search on all studies included at the full-text screening stage through backwards and forward citation tracking using Scopus. No language restriction was applied.

G.B. and M.M. independently assessed the eligibility of all studies retrieved through the database and manual searches and then independently conducted the data extraction and quality assessment. Any conflicts during all the above evaluations were resolved by discussion with a senior author (J.S., and A.H.).

Following discussion with an expert statistician (L.M.), and in accordance with Chapter V of the Cochrane Handbook for Systematic Reviews of Interventions (17), we decided that presenting outcomes as reported in the included systematic reviews, without re-analysing or modifying the original analyses, was an appropriate approach in carrying out the umbrella review. A quantitative approach (18), given the limited number of meta-analyses identified, and the substantial differences in terms of the included populations and mental disorders examined, would not have provided additional value beyond what was already highlighted in the original reviews. Instead, we conducted an overview of reviews to synthesize the current body of the evidence presented, consistent with the approach used in other umbrella reviews (13, 14, 19). We summarised the relevant findings descriptively and in tabular form. We extracted and reported prevalence data and presented meta-analyses or subgroup analyses for specific mental disorders stratified by age, sex, and ASD or ADHD comorbidity, when available. We reported the main findings from systematic reviews that provided a quantitative synthesis of prevalence estimates, using data from the random-effects model when consistent with results from other approaches (e.g., quality-effects model or leave-one-out analysis), when applied. Prevalence estimates were reported with the same decimal precision as in the original reviews. We compared prevalence estimates for mental disorders with those in the general population (DSM-5-TR diagnoses) and organized them by specific diagnoses.

We distinguished between reviews primarily investigating autistic people with ID, and those focusing on specific subgroups of people with ID (e.g., by setting, syndrome, or level of ID) or specific diagnoses. For each systematic review with a narrative synthesis of results, we reported the range of prevalence estimates for each condition based on at least two relevant primary studies, and described the characteristics of the original systematic review, considering only the primary studies relevant to our scope. For every primary study listed in these reviews, a mean IQ cutoff of ≤70 or below was applied to determine ascertainment of ID when the diagnosis was not otherwise specified. We assessed the overall reporting quality of all included reviews using “A Measurement Tool to Assess Systematic Reviews” (AMSTAR-2 tool) (20). Data extraction included: study design, year of publication, number of primary studies (k), corresponding author’s country, timeframe, databases searched, total sample size and range of populations, population characteristics, assessment of mental comorbidity, and key findings relevant to the review objectives, including the type of prevalence investigated. We reported information on N, age, level of ID and whether the considered population was predominantly male or female.

### Quality Appraisal

AMSTAR-2 consists of 16 items, of which 7 (2, 4, 7, 9, 11, 13 and 15) are identified as critical. Each of the items was given a rating between ‘*yes*’, ‘*partial yes*’ and ‘*no*’, which contributes to confidence in the overall quality rating of each included systematic review (20). We considered the critical weakness related to the absence of a pre-registered protocol as not applicable to reviews published in or before 2011, i.e. prior to the introduction of PROSPERO.

## RESULTS

Table 1 shows the main characteristics of the included systematic reviews, whilst Table 2 and Table 3 report the main findings from systematic reviews with a meta-analysis or a subgroup analysis, and from systematic reviews with a narrative synthesis of results, respectively. Of the 26 reviews, seven included a meta-analysis (7, 21–26), and two reported prevalence estimates based on subgroup analyses, or group comparisons (27, 28). These nine reported data on a total of 248,606 people with ID (7, 21–28), including 15,221 people with syndromic ID (23, 24, 26). Systematic reviews with narrative results included a cumulative total of 238,636 people with ID. These figures represent the unadjusted sum of participants across included reviews and may include overlapping primary studies.

**Figure 1.**
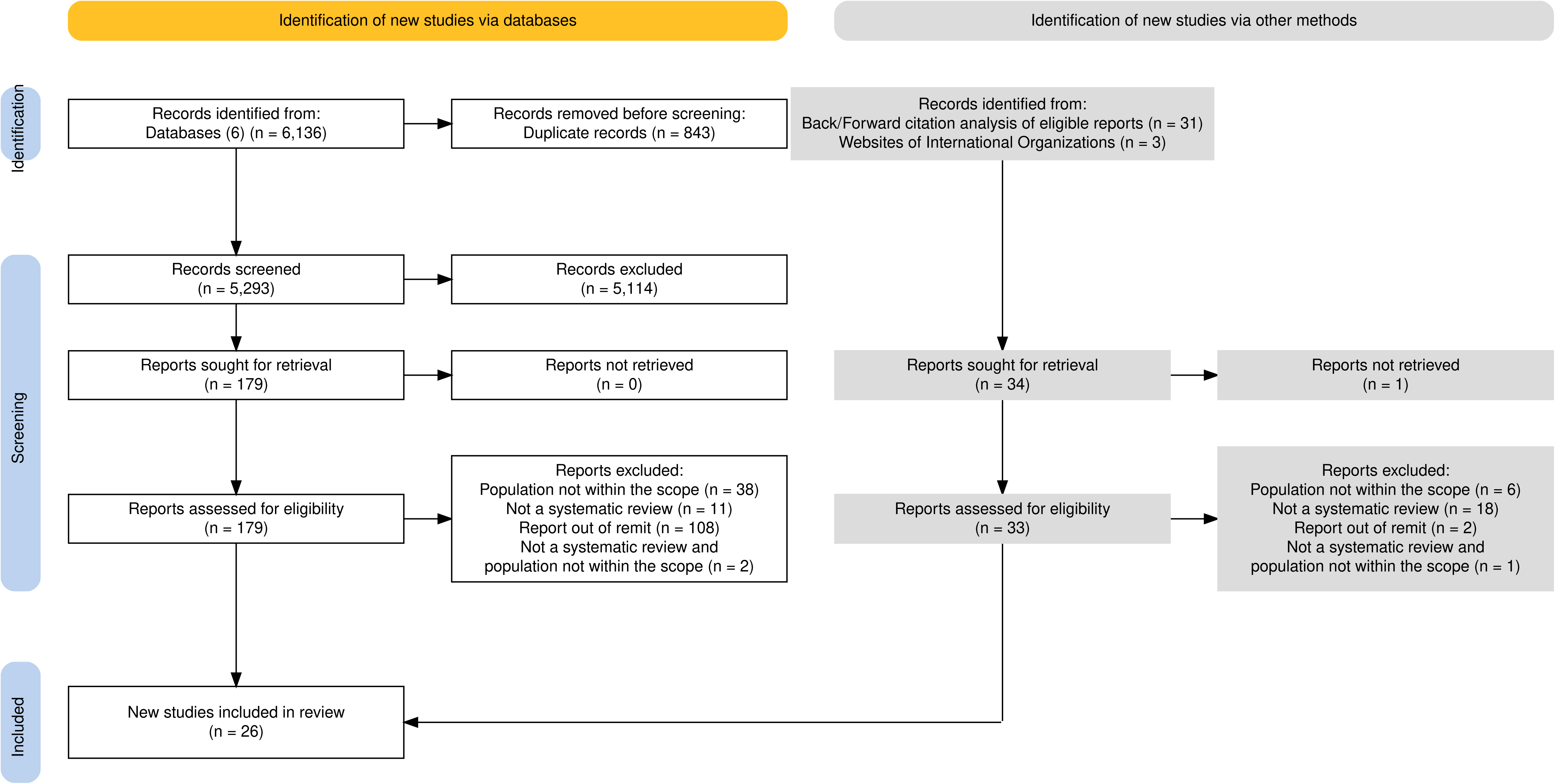
summarises the process of study identification. After EndNote deduplication, database search yielded 5,293 records for title and abstract screening; 179 were included in full-text screening, and 20 reviews met eligibility criteria. Manual search resulted in 34 new reports; one full text was unavailable, and six reviews were included. Supplementary Tables 5-8 present the lists of studies assessed at full-text screening, including those retained and those excluded with corresponding reasons for the decision to include or exclude. Most of the excluded studies were either out of scope, not systematic reviews, or involving non-relevant populations.

**Table 1.** Characteristics of the included review.

| Table 1 - Characteristics of the included reviews |  |  |  |  |
| --- | --- | --- | --- | --- |
| Authors, year, country, topic | Participants | Population characteristics | Mental disorders evaluation | Quality Assessment (AMSTAR-2) |
| Systematic Reviews with a meta-analysis or subgroup-analysis |  |  |  |  |
| (Aman et al 2016, Canada)<sup>21</sup><br>Non-affective psychosis in people with intellectual disabilities | N = 140,189 (52-45,681) | ≥14 yrs<br>All levels of ID<br>M>F<br>Community samples<br>Clinical samples | Clinical/standardized diagnosis | Critical Low |
| (Daveney et al 2019, UK)<sup>22</sup><br>PTSD in people with intellectual disabilities | N = 1,453 (5-1,023) | All age groups<br>All levels of ID (including BIF)<br>F>M<br>Clinical samples | DSM, validated tools | Critical Low |
| (Edwards et al 2022, UK)<sup>23</sup><br>Anxiety disorders in syndromic intellectual disabilities | N = 13,708 (eight ID-related syndromes)<br>22q11.2 deletion syndrome<br>N = 2,272 (range 12-260)<br>Fragile X syndrome<br>N = 3,882 (range 13-1,235)<br>Down syndrome<br>N = 2,152 (range 44-4,975) | Fragile X<br>Age: 0.4-82yrs<br>M/F: 3,461/651<br>22q11.2 deletion syndrome<br>Age: 1-67yrs<br>M/F: 1,189/1,072<br>Down syndrome<br>Age: 1-79yrs<br>M/F: 1,195/1,046 | Validated tools<br>DSM/ICD criteria<br>Medical records | Low |
| (Glasson et al 2020, Australia)<sup>24, a</sup> | 22q11.2 deletion syndrome<br>N = 909 (range 35-802) | 22q11.2 deletion syndrome<br>Age range: 6-17yrs | Clinical assessment<br>Validated tools | Critical Low |
| **Mental disorders in children and adolescents with syndromic intellectual disabilities** | Fragile X syndrome<br>N = 138 (range 31-58)<br>Williams syndrome<br>N = 75 (range 30-45) | M>F<br>Fragile X syndrome<br>Age range: 5-33yrs<br>M>F<br>Williams syndrome<br>Age range: 6-17yrs<br>M = 37, F = 38 |  |  |
| **(Hollocks et al 2019, UK)<sup>27</sup>**<br>**Anxious and depressive disorders in autistic adults with intellectual disabilities** | Anxiety<br>N = 410 (range 13-150)<br>Depression<br>N = 579 (range 13-164) | Anxiety<br>Age range: 16-84yrs<br>M>F<br>Community and inpatient<br>Depression<br>Age range: 16-84yrs<br>M>F<br>Community samples | Clinical assessment<br>Validated tools | Critical Low |
| **(Maiano et al 2018, Canada)<sup>25</sup>**<br>**Anxious and depressive disorders in youth with intellectual disabilities** | N = 57,971 (range 30-43,738) | Age: 0-22yrs<br>ID level: borderline-to profound<br>M>F<br>Community samples<br>Clinical samples<br>Administrative records | Clinical assessment<br>Validated tools | Low |
| **(Mazza et al 2018, Italy)<sup>7</sup>**<br>**Mental disorders in people with intellectual disabilities** | N = 29,958 (range 90-13,631) | Age: > 12yrs<br>Level of ID: mild to profound<br>M>F<br>Community samples<br>Clinical samples | Clinical assessment<br>Medical records | Low |
| **(Oeseburg et al 2011, The Netherlands)<sup>28</sup>**<br>**Mental disorders in people with intellectual disabilities** | N = 2,825 (range 87-1,026) | Age: 0-21yrs<br>Level of ID: borderline-profound<br>Community samples<br>Clinical samples | DSM/ICD criteria<br>Medical records | Critical Low |
| **(Royston et al 2017, UK)<sup>26, b</sup>**<br>**Anxiety disorders in people with Williams syndrome and intellectual disabilities** | N = 391 (10-214)<br>F>M | Mean age: 16.5 yrs (SD = 8.9, range 4-55)<br>IQ mean: 64.4 (SD = 6.3, range 56.6-75.6)<br>F>M<br>Community samples<br>Clinical samples<br>Administrative samples | DSM/ICD criteria | Low |
| **Narrative Reviews** |  |  |  |  |
| **Autistic people with Intellectual Disabilities** |  |  |  |  |
| **(Curnow et al 2023, UK)<sup>35</sup>**<br>**Mental disorders in autistic adults with intellectual disabilities** | N = 4,208 (range 20- 4,059) | Age: 13+<br>M>F<br>Clinical samples<br>Community samples<br>Administrative samples | Medical records | Moderate |
| **(De Giorgi et al 2019, Italy)<sup>37</sup>**<br>**Non-affective psychosis in autistic people with intellectual disabilities** | N = 359 (40-120) | Age: 2-57yrs<br>All levels of ID<br>M>F<br>Clinical samples | Clinical assessment | Low |
| **(Lugo-Marín et al 2019, Spain)<sup>33</sup>**<br>**Psychiatric disorders in** | N = 1,322 (range 23-150) | Mean age: 19.5-40.6yrs<br>M>F<br>Clinical samples | DSM/ICD criteria | Low |
| autistic adults with intellectual disabilities |  | Community samples<br>Mixed |  |  |
| (van Steensel et al 2011, The Netherlands)<sup>31</sup><br>Anxiety disorders in autistic youth with intellectual disabilities | N = 369 (range 12-171) children and adolescents with ASD, PDD-NOS or Asperger's (N=22) and intellectual disabilities | Mean age range: 8.2-16.3yrs<br>Mean IQ range: <40-68 | DSM criteria<br>Validated tools | Low |
| (Varcin et al 2022, Australia)<sup>34</sup><br>Psychosis and bipolar disorder in autistic adults with intellectual disabilities | *Psychosis/Schizophrenia Spectrum Disorders*<br>N = 1,107 (range 23-162)<br>*Bipolar Disorder/Expansive Mood*<br>N = 437 (range 16-162) | *Psychosis/Schizophrenia Spectrum Disorders*<br>Mean age: 19.5-48 yrs<br>M>F<br>*Bipolar Disorder/Expansive Mood*<br>Mean age: 19.5-48 years<br>M>F | DSM/ICD criteria<br>Validated tools<br>Clinical assessment | Low |
| By subgroup (e.g. setting, syndrome, level of intellectual disability) or specific disorder |  |  |  |  |
| (Alexander et al 2003, UK)<sup>42</sup><br>Personality disorders in learning disabilities | NR | All levels of ID<br>Community samples<br>Clinical samples | DSM/ICD criteria<br>Clinical assessment | Critical Low |
| (Bakken et al 2013, Norway)<sup>32</sup><br>Mental disorders in adults with intellectual disabilities | N = 3,996 (range 16-2,005) | Mean age: ~30s (range 29-49yrs)<br>Level of ID: borderline-profound<br>M>F<br>Clinical samples | Validated tools | Critical Low |
| **(Buckley et al 2020, Australia)<sup>29</sup>**<br>**Psychiatric disorders in 6–21 years in children and youth with intellectual disabilities** | N=530 (range 41-178) | Age: 8-21 yrs (children, and adolescents)<br>Level of ID: mild-severe/profound<br>M>F<br>Administrative samples | Clinical assessment | Low |
| **(Huxley et al 2019, UK)<sup>36</sup>**<br>**Substance use disorders in people with intellectual disabilities** | Illicit drug use<br>N = 2,260 (range 30-2,200) | Illicit drug use<br>Mean age: 32yrs<br>Level of ID: borderline-profound<br>M>F<br>Administrative samples<br>Community samples<br>Forensic ID samples | Clinical assessment<br>Validated tools<br>Medical records | Critical Low |
| **(Mevisen et al 2010, The Netherlands)<sup>38</sup>**<br>**PTSD in people with intellectual disabilities** | N = 359 (range 6-310) | Age: children, adolescents, and adults<br>Level of ID: from mild to severe<br>F>M<br>Clinical samples<br>Community samples | DSM criteria<br>Validated tools<br>Medical records | Critical Low |
| **(Rayner et al 2015, UK)<sup>43</sup>**<br>**Personality disorder in offenders with intellectual disabilities** | N = 372 (range 44-164) | Forensic ID samples | Validated tools<br>DSM criteria | Critical Low |
| **(Reardon et al 2015, Australia)<sup>30</sup>** | N = 1,801 (range 74-641) | Age: children/adolescents (range: 5-20yrs) | DSM/ICD criteria<br>Clinical assessment | Critical Low |
| **Anxiety disorders in children/adolescents with intellectual disabilities** |  | Administrative samples<br>Community samples |  |  |
| **(Torr 2008, Australia)<sup>44</sup>**<br>**Personality disorder in offenders with learning disabilities** | N = 769 (range 17–212) | Level of ID: normal- moderate<br>M>F<br>Forensic ID samples | DSM criteria<br>Medical records | Critical Low |
| **(van Duijvenbode et al 2019, The Netherlands)<sup>39</sup>**<br>**Substance use disorder in mild to borderline intellectual disabilities** | People with mild intellectual disability/BIF | 17 studies on children/adolescents (11-21 yrs), 5 on juvenile offenders, 40 on adults (18yrs+), 15 on offenders, 12 on adolescents & adults | Medical records<br>Clinical assessment | Critical Low |
| **(Walton et al 2015, UK)<sup>40, c</sup>**<br>**Unipolar depression in Down syndrome** | N = 1,928 (range 52-497) | Age range: 6-78 yrs<br>Community samples<br>Clinical samples | DSM/ICD criteria | Critical Low |
| **(Walton et al 2016, UK)<sup>45</sup>**<br>**Depression in severe/profound intellectual disabilities** | N = 542 (range 40-184) | People with severe or profound ID<br>Community samples<br>Clinical samples<br>Mixed samples | DSM criteria<br>Validated tools<br>Clinical assessment | Critical Low |
| **(Whitaker et al 2006, UK)<sup>41</sup>** | N = 218,277 (range 90-89,419) | Age: children, adolescents, and adults<br>Level of ID: not specified for all samples | Clinical assessment<br>Validated tools | Critical Low |
| Psychiatric disorders in people with intellectual disabilities |  | Community samples<br>Administrative samples |  |  |

**Abbreviations:** AMSTAR-2: A MeaSurement Tool to Assess Systematic Reviews - Version 2, BIF: Borderline Intellectual Functioning, DSM: Diagnostic and Statistical Manual of Mental Disorders, ICD: International Classification of Diseases, ID: Intellectual Disabilities, IQ: intelligence quotient, PDD-NOS: pervasive developmental disorder - not otherwise specified.
**Notes:** <sup>a</sup> Total population number calculated based on Table 1 from the included review by Glasson et al. <sup>b</sup> Prevalence estimates are reported according to the random effects model. For estimates based on the quality effects model, see the original review by Royston et al. <sup>c</sup> The sample size (N) was calculated using data from Table 1 of the original review by Walton et al., specifically the column 'Frequency of depression in sample,' and by considering the T2 sample size for the two included longitudinal studies.

**Table 2.** Prevalence of mental disorders in people with intellectual disabilities: evidence from systematic reviews with meta-analyses or subgroup analyses.

|  |  |  |
| --- | --- | --- |
| Mental disorders co-occurrence | Findings (prevalence rate, 95% CI, N = population, k= number of primary studies relevant to intellectual disabilities in the included reviews) | Authors |
| Schizophrenia Spectrum and Other Psychotic Disorders | All psychotic disorders prevalence: 3.46%, [95% CI: 2.51%-4.42%], k=25;<br>Psychosis point prevalence: 3.75%, [95% CI 2.63%-4.87%], N=2,664, k=11;<br>Psychosis undefined prevalence: 3.30%, [95% CI: 2.02%-4.59%], N=136,538, k=11;<br>All psychotic disorders (high quality studies in community samples) prevalence: 2.91%, [95% CI 1.38%-4.44%], k=5;<br>Schizophrenia prevalence: 3.55%, [95% CI: 2.66%-4.44%], k=17.<br>*Subgroup Analyses by sex (k=9) and level of intellectual disability level (k=7)*<br>Prevalence of psychosis in males: 3.78%, [95% CI: 2.56%-5.00%], N=28,830;<br>Prevalence of psychosis in females: 4.10%, [95% CI: 2.86%-5.35%], N=22,907;<br>Prevalence of psychosis in mild ID: 5.55%, [95% CI: 3.86%-7.25%], N=7,160;<br>Prevalence of psychosis moderate ID: 4.21%, [95% CI: 2.56%-5.86%], N=4,231;<br>Prevalence of psychosis in severe ID: 0.89%, [95% CI: 0.15%-1.62%], N=3,274. | (Aman et al)<sup>21</sup> |
|  | Schizophrenia prevalence: 4.8%, [95% CI: 2.4%-9.1%], N=21,125, k=9;<br>Unspecified psychotic disorders prevalence: 3.9%, [95% CI: 0.8%-16.8%], N=20,885, k=8. | (Mazza et al)<sup>7</sup> |
| Mood Disorder |  |  |
|  | Mood disorder prevalence: 6.7%, [95% CI: 5.0%-8.8%], N=9,118, k=12. | (Mazza et al)<sup>9</sup> |
| Depressive Disorder |  |  |
|  | Current depression prevalence: ASD+ID: 14%, [95% CI 5%-28%], N=58/512, k=6, Vs ASD-only: 26%, [95% CI 20%-32%], N=326/1,413, k=16. | (Hollocks et al)<sup>27</sup> |
|  | Combined depressive disorder prevalence: 2.8%, [95% CI: 1.1%-6.7%], N=55,090, k=10;<br>Dysthymic disorder prevalence: 3.4%, [95% CI: 1.5%-7.4%], k=3;<br>Major depressive disorder prevalence: 2.5%, [95% CI: 1.2%-5.4%], k=4. | (Maïano et al)<sup>25</sup> |
|  | *Subgroup Analysis by age and level of intellectual disability:*<br>Combined depressive disorder prevalence in adolescents: 1.4%, [95% CI: 0.4%-5.1%], N=43,962, k=3;<br>Borderline: 17.1%, [95% CI 8.4%-31.8%], k=2;<br>Mild: 9.4%, [95% CI 3.8%-21.6%], k=5;<br>Moderate: 3.5%, [95% CI 1.4%-8.5%], k=3;<br>Severe: 2.6%, [95% CI 0.6%-10.3%], k=3;<br>Profound: 4%, [95% CI 0.6%-25.6%], k=2;<br>Unspecified: 3.4%, [95% CI 0.7%-15.2%], k=2. |  |
| **Obsessive compulsive disorder** |  |  |
|  | OCD prevalence: ASD+ID: 24%, [95% CI 14%-36%], N=55/177, k=3 Vs ASD without ID: 20%, [95% CI 10%-34%], N=210/970, k=7. | (Hollocks et al)<sup>27</sup> |
|  | OCD prevalence: 2.4%, [95% CI: 0.8%-7.2%], N=1,664, k=6.<br>*Subgroup Analysis by age and level of intellectual disability*<br>Adolescents: 12.0%, [95% CI: 7.2%-19.2%], k=2.<br>Borderline: 3.3%, [95% CI 0.5%-20.3%], k=2;<br>Mild: 2.5%, [95% CI 0.8%-7.6%], k=2;<br>Moderate: 1.6%, [95% CI 0.3%-8.4%], k=2. | (Maïano et al)<sup>25</sup> |
| **Anxiety Disorder** |  |  |
|  | Current anxiety prevalence: ASD+ID: 20%, [95% CI 7%-39%], N=79/394, k=6 Vs ASD-only: 24%, [95% CI 19%-43%], N=352/1,050, k=7. | (Hollocks et al)<sup>27</sup> |
|  | Combined anxiety disorder prevalence: 5.4%, [95% CI: 2.5%-11.5%], N=57,774, k=18;<br>AGO prevalence: 0.6%, [95% CI: 0.1%-3.6%], k=2;<br>PD prevalence: 0.3%, [95% CI: 0.1%-1.2%], k=2;<br>GAD prevalence: 2.2%, [95% CI: 0.5%-9.1%], k=5;<br>SAD prevalence: 5.0%, [95% CI: 1.8%-13.3%], k=4;<br>SP prevalence: 2.7%, [95% CI: 1.2%-5.8%], k=5; | (Maïano et al)<sup>25, a</sup> |
|  | SPHOBIA prevalence: 11.5%, [95% CI: 4.0%-28.9%], k=4.<br>*Subgroup Analysis by age (anxiety disorders)*<br>Combined anxiety disorder prevalence in children: 1.2%, [95% CI: 0.02%-49.1%], N=43,894, k=3;<br>Combined anxiety disorder prevalence in adolescents: 7.9%, [95% CI: 0.6%-54.6%], N=44,079, k=5;<br>GAD prevalence in adolescents: 8.7%, [95% CI: 2.4%-27.1%], k=2;<br>SP prevalence in adolescents: 5.4%, [95% CI: 1.9%-14.6%], k=2;<br>SPHOBIA prevalence in adolescents: 19.8%, [95% CI: 11.8%-31.3%], k=2.<br>*Subgroup Analyses by level of intellectual disability (anxiety disorders)*<br>Borderline: 3.3%, [95% CI 0.5%-20.3%], k=2;<br>Mild: 8.1%, [95% CI 5.2%-12.3%], k=4;<br>Moderate: 1.7%, [95% CI 0.4%-6.7%], k=2;<br>Severe: 3.5%, [95% CI 1.3%-8.9%], k=2. |  |
|  | Combined anxiety disorder prevalence: 5.5%, [95% CI: 3.3%-9%], N=4,494, k=9 | (Mazza et al.)<sup>7</sup> |
|  | Any anxiety disorder prevalence: 17.1%, [95% CI: 15.1-19.4%], N=1,202, k=3. | (Oeseburg et al)<sup>28</sup> |
| **Post-Traumatic Stress Disorder** |  |  |
|  | PTSD: 10.0%, [95% CI: 0.4%-19.5%], k=4.<br>*Subgroup Analyses by age (k=2):*<br>PTSD prevalence in adults: 8.0%, [95% CI: -8%-24%], N=1,333, k=2<br>PTSD prevalence in children: 12.95%, [95% CI: -8.2%-33%], N=120, k=2. | (Daveney et al)<sup>22</sup> |
|  | PTSD prevalence: 1.1%, [95% CI: 0.3%-3.7%], k=4. | (Maïano et al)<sup>25</sup> |
| **Personality, Disruptive, Impulse-Control, and Conduct Disorder** |  |  |
|  | Personality disorder prevalence: 2.8%, [95% CI: 1.2%-6.8%], N=3,949, k=5. | (Mazza et al)<sup>7</sup> |
|  | Conduct disorder prevalence: 5.1%, [95% CI: 4.1%-6.4%], N=1,535, k=5;<br>Oppositional defiant disorder prevalence: 12.4%, [95% CI: 10.7%-14.4%], N=1,202 k=3. | (Oeseburg et al)<sup>28</sup> |
| **Mental disorders in syndromic intellectual disabilities** |  |  |
|  | *Prevalence of anxiety disorders in:*<br>Down syndrome: 8%, [95% CI: 4%-13%], N=2,152, k=11;<br>Fragile X syndrome: 49%, [95% CI: 39%-60%], N=3,882, k=19;<br>Males with Fragile X syndrome: 43%, [95% CI: 23%-62%], N=1,124, k=7;<br>22q11.2 deletion syndrome: 38%, [95% CI: 32%-45%], N=2,272, k=34.<br>*Prevalence of specific anxiety disorders in:*<br>Down syndrome<br>SPHOBIA/phobic anxiety: 4%, [95% CI: 0%-9%], N=933, k=4;<br>OCD: 2%, [95% CI: 0%-4%], N=933, k=4;<br>Selective mutism: 6%, [95% CI: 2%-10%], N=155, k=2.<br>Fragile X syndrome<br>SPHOBIA: 52%, [95% CI: 38%-67%], N=44, k=2;<br>Social anxiety: 28%, [95% CI: 16%-41%], N=123, k=7;<br>GAD: 14%, [95% CI: 5%-22%], N=172, k=5;<br>SAD: 13%, [95% CI: 7%-19%], N=125, k=5;<br>PD/AGO: 27%, [95% CI: -12%-65%], N=32, k=2;<br>OCD: 16%, [95% CI: 2%-30%], N=187, k=4.<br>22q11.2 deletion syndrome<br>SPHOBIA: 28%, [95% CI: 21%-35%], N=1,412, k=18;<br>Social anxiety/social phobia: 9%, [95% CI: 7%-11%], N=1,252, k=15;<br>GAD: 13%, [95% CI: 10%-16%], N=1,448, k=17;<br>SAD: 6%, [95% CI: 4%-8%], N=762, k=14;<br>OCD: 9%, [95% CI: 7%-12%], N=1,541, k=21;<br>PD: 1%, [95% CI: 0%-2%], N=449, k=4;<br>AGO: 2%, [95% CI: 0%-3%], N=226, k=3;<br>PTSD: 2%, [95% CI: 1%-3%], N=483, k=6. | (Edwards et al)<sup>23, b</sup> |
|  | Fragile X syndrome<br>Any anxiety disorder prevalence: 42%, [95% CI: 35%-48%], N=107, k=2.<br>Williams syndrome<br>Any anxiety disorder prevalence: 46%, [95% CI: 35%-58%], N=75, k=2.<br>22q11.2 deletion syndrome<br>Any anxiety disorder prevalence: 38%, [95% CI: 27%-50%], N=909, k=3. | (Glasson et al)<sup>24</sup> |
|  | Williams syndrome<br>Any anxiety disorder prevalence: 48%, [95% CI: 30%-67%], n=391 k=7;<br>SPHOBIA prevalence: 40%, [95% CI: 27%-54%], k=9;<br>GAD prevalence: 11%, [95% CI: 5%-19%], k=7;<br>SAD prevalence: 7%, [95% CI: 2%-15%], k=6;<br>Social anxiety disorder prevalence: 1%, [95% CI: 0%-3%], k=6;<br>PD prevalence: 2%, [95% CI: 1%-4%], k=6;<br>PTSD prevalence: 2%, [95% CI: 0%-4%], k=4;<br>AGO prevalence: 2%, [95% CI: 0%-5%], k=3;<br>OCD prevalence: 4%, [95% CI: 2%-6%], k=7. | (Royston et al)<sup>26, c</sup> |

**Abbreviations:** AGO: agoraphobia, ASD: autism spectrum disorder, GAD: generalized anxiety disorder, ID: intellectual disabilities, OCD: obsessive-compulsive disorder, PD: panic disorder, PTSD: post-traumatic stress disorder, SAD: separation anxiety disorder, SP: social phobia, SPHOBIA: specific phobia.
**Notes:** <sup>a</sup> According to the original review by Mañano et al., the combined prevalence estimates for anxiety disorders included estimates for OCD and PTSD. <sup>b</sup> For prevalence estimates calculated using the quality effects model and the leave-one-out analysis, please refer to the original systematic review by Edwards et al. <sup>c</sup> Prevalence estimates reported according to the random effects model. For estimates based on the quality effects model, please see the original review by Royston et al. (Supplementary Materials). According to the original review by Royston et al., the combined prevalence estimates for anxiety disorders included estimates for OCD and PTSD.

**Table 3.** Prevalence of mental disorders in people with intellectual disabilities: evidence from narrative reviews.

|  |  |  |
| --- | --- | --- |
|  | Findings (k= number of primary studies relevant to intellectual disabilities included in the review) | Authors |
| **Autistic people with Intellectual Disabilities** |  |  |
|  | Anxiety lifetime prevalence range: 17%-53%, k=2;<br>Anxiety current prevalence range: 40%-50%, k=2;<br>Depression lifetime prevalence range: 13%-24%, k=2;<br>Psychosis lifetime prevalence range: 10%-17%, k=2. | (Curnow et al)<sup>35</sup> |
|  | Non affective psychosis prevalence range: 0%-25.1%, k=5. | (De Giorgi et al)<sup>37</sup> |
|  | SUD prevalence range: 0%-11.9%, k=4;<br>Schizophrenia spectrum disorder prevalence range: 1.3%-26.1%, k=10;<br>Mood disorder prevalence range: 3.4%-36.1%, k=5;<br>Anxiety disorders prevalence range: 0%-32.7%, k=7;<br>Personality disorders prevalence range: 0%-36.1%, k=6. | (Lugo-Marín et al)<sup>33</sup> |
|  | Prevalence range of any anxiety disorder or impairing anxiety level above clinical cut off: 41.7%-42.7%, k=3;<br>GAD prevalence range: 9.4%-24.6%, k=2;<br>Social anxiety disorder prevalence range: 16.4%-19.9%, k=2;<br>SAD prevalence range: 10.5%-14.8%, k=2;<br>SPHOBIA prevalence range: 17%-67.2%, k=3. | (van Steensel et al)<sup>31</sup> |
|  | Psychosis/Psychotic disorder/Schizophrenia Spectrum disorder prevalence range: 0%-32.6%, k=10;<br>Bipolar disorder (I, II, NOS, expansive mood) prevalence range: 3.6%-6.2%, k=4. | (Varcin et al)<sup>34</sup> |
| **By subgroup (e.g. setting, syndrome, level of intellectual disability) or specific disorder** |  |  |
|  | Personality disorders prevalence range: <1%-91% in community settings, 22%-92% in hospital-based populations, (k=14). | (Alexander et al)<sup>42</sup> |
|  | *Inpatient units:*<br>Psychosis (schizophrenia, psychosis, psychotic disorders, and affective psychosis) prevalence range: 6%-59%, k=19; | (Bakken et al)<sup>32, a</sup> |
|  | Affective disorders (depression, mood disorder, bipolar disorder, affective disorder, and mania) prevalence range: 9%-42%, k=19;<br>Anxiety disorders (anxiety and OCD) prevalence range: 0%-8% (outlier excluded), k=18;<br>Other mental illness disorder (eating disorders, organic disorders, substance-related disorders, personality disorders, impulse control disorders, and dementia) prevalence range: from 0% to >63%, k=17. |  |
|  | *Children:*<br>Depressive disorders prevalence range: 3%-5%, k=3;<br>Conduct disorders prevalence range: 3%-21%, k=4;<br>Any anxiety disorder prevalence range: 7%-34%, k=3. | (Buckley et al)<sup>29</sup> |
|  | *22q11.2 Deletion Syndrome*<br>Oppositional defiant disorders range: 10%-14%, n=909, k=3;<br>Mood disorders range: 3%-7%, n=909, k=3;<br>Psychotic disorders range: 0%-5%, n=909, k=3;<br>Conduct disorders range: 0%, n=837, k=2. | (Glasson et al)<sup>24</sup> |
|  | SUDs (illicit drugs) prevalence range: 1.7%-3%, N=2,260, k=2. | (Huxley et al)<sup>36</sup> |
|  | PTSD prevalence range: 2.5%-60%, k=3. | (Mevisen et al)<sup>38</sup> |
|  | Any mood disorder prevalence range: 4.4%-14.0%, k=2;<br>Eating disorders prevalence range: 0.2%-3.4%, k=2. | (Oeseburg, et al)<sup>28</sup> |
|  | *Offenders:*<br>Any personality disorder prevalence range: 9.8%-80%, k=3;<br>Antisocial or borderline disorders prevalence range: 22.1%-33%, k=2;<br>Personality disorders not otherwise specified prevalence range: 9.8%, k=2. | (Rayner et al)<sup>43, b</sup> |
|  | *Children:*<br>Anxiety disorder prevalence range: 3%-21.9%, k=6;<br>SAD prevalence range: 2.1%-17.6%, k=5;<br>Social anxiety disorder prevalence range: 0.8%-10.8%, k=6;<br>GAD prevalence range: 0%-5.4%, k=4;<br>SPHOBIA (k=3), panic disorder (k=3), agoraphobia (k=3) prevalence range: mostly ≤2% (one outlier for SPHOBIA | (Reardon et al)<sup>30</sup> |
|  | excluded). |  |
|  | *Offenders:*<br>Any personality disorder prevalence range: 12%-54.8%, k=8;<br>Antisocial personality disorders prevalence range: 13%-38%, k=3. | (Torr)<sup>44, c</sup> |
|  | Adolescents: SUD prevalence range: 0.1-2.7%;<br>Adults: SUD prevalence range: 0.5-46.0%. | (van Duijvenbode et al)<sup>39</sup> |
|  | Unipolar depression incidence range: 5%-13%, k=8. | (Walton et al)<sup>40</sup> |
|  | Unipolar depression prevalence range: 1.1%-59%, k=5. | (Walton et al)<sup>45</sup> |
|  | *Children, k=6:*<br>Depression prevalence range: 1.5%-11%;<br>Anxiety disorder prevalence range: 8.7%-21.9%;<br>Conduct disorder prevalence range: 3%-25%.<br>*Children and Adults, k=12:*<br>Any psychotic disorder prevalence range: 0%-10%;<br>Schizophrenia disorder prevalence range: 0.4%-6.2%;<br>Depression prevalence range: 0%-6%;<br>Affective disorders prevalence range: 0%-2.6%;<br>Personality disorders current prevalence range: 0.2%-3.9%;<br>Anxiety disorders prevalence range: 0.1%-5.2% | (Whitaker et al)<sup>41</sup> |

**Abbreviations:** GAD: generalized anxiety disorder, OCD: obsessive-compulsive disorder, PTSD: post-traumatic stress disorder, SAD: separation anxiety disorder, SP: social phobia, SPHOBIA: specific phobia, SUD: substance use disorder. **Notes** <sup>a</sup> Prevalence ranges were extracted from Table 1 of the original systematic review by Bakken et al. (2013). <sup>b</sup> The original review by Rayner et al included two studies that used overlapping data. <sup>c</sup> Prevalence ranges were extracted from Table 1 of the original systematic review by Torr et al. (2013).

Six systematic reviews focused on children/adolescents or youth up to 22 years (24, 25, 28–31), three on adults (32–34), five on adolescents/adults (7, 21, 27, 35, 36), eight across all ages (22, 23, 26, 37–41) and four with limited or no age data (42–45). Most of the studies included predominantly male participants (7, 21, 23–25, 27, 29, 32–37, 44), while nine reported limited or no sex-related information (28, 30, 31, 39–43, 45). Only three studies reported populations with an overall female predominance (22, 26, 38).

More detailed information on characteristics and diagnostic procedures was available in reviews with meta-analyses. Most of the reviews reported multiple sites and strategies of recruiting study participants and included primary studies in which diagnoses of mental disorders were established through structured or semi-structured clinical interviews, based on international diagnostic criteria or other validated assessment tools (7, 21–31, 33, 34, 36, 38, 40, 41, 44, 45).

Eleven reviews were conducted in the UK (22, 23, 26, 27, 35, 36, 40–43, 45), five in Australia (24, 29, 30, 34, 44), four in the Netherlands (28, 31, 38, 39), two in Canada (21, 25), two in Italy (7, 37), one each in Norway (32), and Spain (33). Two reviews are included in both Tables 2 and 3, as they reported meta-analyses or subgroup analyses alongside a narrative synthesis of prevalence data from individual studies on different mental disorders (24, 28). AMSTAR-2 ratings were *critically low* for 16 reviews (21, 22, 24, 27, 28, 30, 32, 36, 38–45), *low* for nine (7, 23, 25, 26, 29, 31, 33, 34, 37), and *moderate* for one (35). Eighteen reviews used a quality assessment tool for the included primary studies (7, 21–29, 31, 33–37, 40, 45). The most frequent critical weaknesses found were the absence of a pre-established review protocol, and the lack of a list of excluded studies. Supplementary Tables 2-4 provide scoring details and a description of the AMSTAR-2 tool.

## Mental disorders in people with intellectual disabilities

### Schizophrenia Spectrum and Other Psychotic Disorders

In adolescents and adults with ID, Aman et al. (21) reported a prevalence of 3.46% (95% CI: 2.51%-4.42%) for all psychotic disorders in 140,189 participants (≥14 years, k=25), slightly lower than Mazza et al. who reported 3.9% (95% CI: 0.8%-16.8%, k=8) for unspecified psychotic disorder in 20,885 participants (>12 years) (7).

Aman et al. found a 2.91% (95% CI: 1.38%-4.44%) point prevalence including five high-quality studies for all psychotic disorders, 3.75% (95% CI: 2.63%-4.78%, N=2,664, k=11) point prevalence for psychosis, and 3.30% (95% CI: 2.02%-4.59%, N=136,538, k=11) in studies without a clear prevalence type (21). While they estimated a prevalence of 3.55% (95% CI: 2.66%-4.44%, k=17) for schizophrenia, Mazza et al. highlighted a prevalence of schizophrenia of 4.8% (95% CI: 2.4%-9.1%, k=9) in 21,125 adolescents and adults with ID (7, 21). With reference to sex, sensitivity analyses by Aman et al. (21) showed the prevalence of psychosis being 3.78% (95% CI: 2.56%-5.00%, N=28,830) in males and of 4.10% (95% CI: 2.86%-5.35%, N=22,907) in females (k=9) and by level of ID, a prevalence of psychosis of 5.55% (95% CI: 3.86%-7.25%) in mild, 4.21% (95% CI: 2.56%-5.86%) in moderate, and 0.89% (95% CI: 0.15%-1.62%) in severe ID (k=7).

Systematic reviews with narrative results also addressed psychotic disorders in people with ID; Whitaker et al. reported prevalence ranges of 0%-10% for any psychotic disorder, and 0.4%-6.2% for schizophrenia across the lifespan (41).

According to DSM-5-TR (1), the lifetime prevalence of schizophrenia spectrum disorders in the general population is estimated at 0.3%-0.7%.

### Mood Disorders

Mazza et al., reported a prevalence of what they generally named “mood disorders” of 6.7% (95% CI: 5.0%-8.8%, k=12) in 9,118 people with ID (7), while Oeseburg et al. reported prevalence ranges of 4.4%-14% (k=2) for any mood disorder (28).

In the general population, the 12-month prevalence of all mood disorders among adults is 9.7% (46).

### 1. Depressive Disorder

Maïano et al. reported a prevalence of combined subtypes of depressive disorder of 2.8% in youth (<21 years) with ID (95% CI: 1.1%-6.7%, N=55,090, k=10), 3.4% (95% CI: 1.5%-7.4%, k=3) for dysthymic disorder) and 2.5% (95% CI: 1.2%-5.4%, k=4) for major depressive disorder (25).

The same study reported a prevalence of depressive disorder in adolescents (12-22 years) with ID of 1.4% (95% CI: 0.4%-5.1%, k=3) (25). By level of ID, the prevalence of depressive disorder was 17.1% (95% CI: 8.4%-31.8%, k=2) in Borderline Intellectual Functioning (BIF), 9.4% (95% CI: 3.8%-21.6%, k=5) in mild, 3.5% (95% CI: 1.4%-8.5%, k=3) in moderate, 2.6% (95% CI: 0.6%-10.3%, k=3) in severe, 4% (95% CI: 0.6%-25.6%, k=2) in profound, and 3.4% (95% CI: 0.7%-15.2%, k=2) in those with unspecified ID (25).

Systematic reviews with narrative results reported variable prevalence rates of depression in children and adolescents with ID. Buckley et al. found prevalence ranges of 3%-5% (k=3) (29). Whitaker et al. reported ranges of 1.5%-11% (N= 218,277, k=6) (41), and in a broader sample including both children and adults (k=12), they reported prevalence ranges of 0%-6% for depression and 0%-2.6% for affective disorders. Regarding cognitive level, Walton et al. found unipolar depression prevalence range of 1.1%-59% (k=5, N= 542) in people with severe or profound ID (45).

Globally, depression affects 3.8% of the population (WHO), including 5.7% of adults older than 60 years; in the U.S., the 12-month prevalence of major depressive disorder is about 7% (DSM-5-TR) (1, 47).

### 2. Bipolar Disorder

We found no meta-analytic prevalence estimates of Bipolar I and II Disorder (BD) in people with ID. Annual prevalence of BD I in the population without ID has been estimated at 0.0%-0.6%, while lifetime prevalence of BD II is around 0.3% (1).

### Obsessive Compulsive Disorder

Maïano et al. reported the prevalence of obsessive-compulsive disorder (OCD) as 2.4% (95% CI: 0.8%-7.2%, N=1,664, k=6) in children/adolescents with ID, and 12% (95% CI: 7.2%-19.2%, k=2) in adolescents with ID (25).

By level of ID, Maïano et al. reported prevalence of OCD as 3.3% (95% CI: 0.5%-20.3%, k=2) in BIF, 2.5% (95% CI: 0.8%-7.6%, k=2) in mild, and 1.6% (95% CI: 0.3%-8.4%, k=2) in moderate ID, while no data were available for other levels of ID (25).

DSM-5-TR estimates the 12-month OCD prevalence of 1.1%-1.8% (1), consistent with the meta-analysis by Fawcett et al. who report the global prevalence of OCD as 1.1% (current), 0.8% (period), and 1.3% (lifetime) (48). Regarding the prevalence of OCD in children and adolescents, Krebs et al.’s review report an estimated prevalence between 0.25%-4% (k=3) (49).

### Anxiety Disorders

We found the combined prevalence of anxiety disorders to be 5.4% (95% CI: 2.5%-11.5%, k=18) among 57,774 children, adolescents, and young adults with ID, and 5.5% (95% CI: 3.3%-9.0%) among 4,494 adolescents and adults with ID (aged over 12 years, k=9) (7, 25).

In adults, Maïano et al. reported prevalence estimates for specific anxiety disorders: Agoraphobia 0.6% (95% CI: 0.1%-3.6%, k=2), Panic Disorder (PD) 0.3% (95% CI: 0.1%-1.2%, k=2), Generalized Anxiety Disorder (GAD) 2.2% (95% CI: 0.5%-9.1%, k=5), Separation Anxiety Disorder (SAD) 5% (95% CI: 1.8%-13.3%) (k=4), Social Phobia (SP) 2.7% (95% CI: 1.2%-5.8%, k=5) and Specific Phobias (SPHOBIA) 11.5% (95% CI: 4.0%-28.9%, k=4) (25).

Among children (4-11 years), the overall prevalence of anxiety disorders was 1.2% (95% CI: 0.02%-49.1%, N=43,894, k=3). For adolescents (12-21 years), the overall prevalence was 7.9% (95% CI: 0.6%-54.6%, N=44,079, k=5), with GAD at 8.7% (95% CI: 2.4%-27.1%, k=2), SP 5.4% (95% CI: 1.9%-14.6%, k=2) and SPHOBIA at 19.8% (95% CI: 11.8%-31.3%, k=2) (25). Subgroup analyses by level of ID showed prevalence estimates of combined anxiety disorder of 3.3% (95% CI: 0.5%-20.3%, k=2) in youth (0-22 years) with BIF, 8.1% (95% CI: 5.2%-12.3%, k=4) in mild, 1.7% (95% CI: 0.4%-6.7%, k=2) in moderate, and 3.5% (95% CI: 1.3%-8.9%, k=2) in severe ID (25).

Oeseburg et al. in their sample of 1,202 children and adolescents (0-21 years) with ID reported an overall prevalence of any anxiety disorder of 17.1% (95% CI: 15.1%-19.4%, k=3) (28).

Systematic reviews with narrative results reported wide variability in prevalence estimates of anxiety disorders in children and adolescents with ID. Buckley et al. found ranges of 7%-34% (k=3) (29), while Reardon et al. reported prevalence of 3%-21.9% for any anxiety disorder (N=1,801, k=6), 2.1%-17.6% (k=5) for SAD, 0.8%-10.8% (k=6) for social anxiety disorder, and 0%-5.4% (k=4) for GAD (30). SP, PD, and agoraphobia were mostly <2% (each k=3) (30) Whitaker et al. reported prevalence of 8.7%-21.9% (k=6) in children and adolescents, and 0.1%-5.2% (k=12) in mixed samples of children and adults (41).

Globally, anxiety disorders affect 4.05% of the population (50) with 31.1% of U.S. adults experiencing any anxiety disorder during the lifetime (51), while the cumulative incidence by 18 years is 7.85% in girls and 4.58% in boys (52).

### Post-Traumatic Stress Disorder

Mevissen et al. reported Post-Traumatic Stress Disorder (PTSD) prevalence rates of 2.5%-60% (k=3) in 359 people with ID across the lifespan (38). These findings were later updated by Daveney et al., which reported a prevalence of 10.0% (95% CI: 0.4%-19.5%, k=4) in 1,453 people with ID across the lifespan with subgroup analyses by age showing PTSD prevalence at 8.0% (95% CI: –8.0%-24.0%, N=1,333, k=2) in adults, and 12.95% (95% CI: –8.2%-33.0%, N=120, k=2) in children and adolescents (22). Maïano et al. reported PTSD prevalence of 1.1% (95% CI: 0.3%-3.7%, k=4) in children and adolescents with ID (25).

Schincariol et al. report an overall PTSD prevalence of 23.95% (95% CI: 20.74%-27.15%) in the general population (53). National lifetime estimates of PTSD vary considerably across country income groups and WHO regions but averaged 3.9% overall (54).

### Personality Disorders, Disruptive, Impulse-Control, and Conduct Disorder

One review estimated the prevalence of personality disorders of 2.8% (95% CI: 1.2%-6.8%, N=3,949, k=5) in adolescents and adults with ID (7). Regarding specific disorders in children and adolescents (<19 years), Oeseburg et al. reported prevalence of Conduct Disorder (CD) at 5.1% (95% CI: 4.1%-6.4%, N=1,535, k=5), and 12.4% (95% CI: 10.7%-14.4%, N=1,202, k=3) for oppositional defiant disorder (28).

Systematic reviews with narrative results also reported on CD in children and adolescents with ID: Buckley et al. found prevalence ranges of 3%-21% (k=4) (29), and Whitaker et al. of 3%-25% (k=6) (41), while for personality disorder in people with ID, Whitaker et al. reported prevalence ranges of 0.2%-3.9% (k=12) in mixed samples of children and adults (41).

In Western countries, the prevalence of any personality disorder in the general population is about 12% (55), while one-year prevalence estimates for CD in high-income countries range from 2% to over 10%, with a median of 4% (1).

### Other mental disorders (Eating, and Substance Use Disorders)

No reviews reported meta-analyses for these mental disorders in people with ID; however, systematic reviews with narrative results have highlighted the following findings. Huxley et al. reported a prevalence range of Substance Use Disorder (SUD) for illicit drugs at 1.7%-3%, in 2,260 people with ID (k=2) (36), and van Duijvenbode et al. found prevalence of SUD at 0.1%-2.7% in adolescents, and 0.5%-46% in adults with mild ID and in BIF (39). Oeseburg et al reported the prevalence range for eating disorders at 0.2%-3.4% (k=2) (28).

Compared with the general population, the lifetime and 12-month prevalence of eating disorders have been estimated at 0.91% and 0.43%, respectively (56). According to the World Drug Report 2025 by the United Nations Office on Drugs and Crime, approximately 6% of the general population used illicit drugs in 2023 (57).

## Mental disorders in syndromic intellectual disabilities

### Down syndrome

Edwards et al. included 11 studies involving 2,152 people with Down syndrome. They reported a prevalence of 8% (95% CI: 4%-13%, k=11) for any anxiety disorder, and 4% (95% CI: 0%-9%, k=4) for SPHOBIA/phobic anxiety in 933 participants. The same sample showed a 2% (95% CI: 0%-4%, k=4) prevalence for OCD, and 6% (95% CI: 2%-10%, k=2) prevalence for selective mutism in 155 people with Down syndrome (23). Walton et al. reported a prevalence of unipolar depression ranging 5%-13% (k=8) in 1,928 people with Down syndrome across the lifespan (age 6-78 years) (40)

### Fragile X syndrome

Edwards et al. reported a prevalence of any anxiety disorder (including PTSD) of 49% (95% CI: 39%-60%, k=19) in 3,882 people with Fragile X syndrome across the lifespan, and 43% (95% CI: 23%-62%, k=7) in the 1,214 male participants (23). Consistent with these findings, Glasson et al. reported a prevalence of any anxiety disorder of 42% (95% CI: 35%-48%, k=2) in 107 children (24).

Regarding specific anxiety disorder in Fragile X syndrome, Edwards et al. reported the following prevalence estimates: SPHOBIA, 52% (95% CI: 38%-67%, k=2); social anxiety, 28% (95% CI: 16%-41%, k=7); GAD, 14% (95% CI: 5%-22%, k=5); SAD, 13% (95% CI: 7%-19%, k=5); PD with agoraphobia, 27% (95% CI: 12%-65%, k=2), and OCD, 16% (95% CI: 2%-30%, k=4) (23).

### Williams syndrome

Glasson et al. reported a prevalence of any anxiety disorder of 46% (95% CI: 35%-58%, k=2) in 75 children and adolescents (24). Royston et al. reported a prevalence of any anxiety disorder of 48% (95% CI: 30%-67%, k=7) in 391 people with Williams syndrome and mild ID; SPHOBIA of 40% (95% CI: 27%-54%, k=9); GAD of 11% (95% CI: 5%-19%; k=7); SAD of 7% (95% CI: 2%-15%, k=6); social anxiety disorder of 1% (95% CI: 0%-3%, k=6); PD of 2% (95% CI: 1%-4%, k=6); PTSD of 2% (95% CI: 0%-4%, k=4); agoraphobia of 2% (95% CI: 0%-5%, k=3); OCD of 4% (95% CI: 2%-6%, k=7) (26). The prevalence estimates using the random-effects analytic model were similar to those according to the quality effects model (26), and PTSD and OCD have been included in the overall estimate of anxiety disorders.

### 22q11.2 deletion syndrome

For any anxiety disorder in the 22q11.2 deletion syndrome, Edwards et al. estimated a prevalence of 38% (95% CI: 32%-45%, N= 2,272, k=34) across the lifespan, equal to the prevalence of any anxiety disorder reported by Glasson et al. (38%, 95% CI: 27%-50%, k=3) in 909 children/adolescents (6-17 years) (23, 24).

For specific anxiety disorder, Edwards et al. reported a prevalence estimate of 28% (95% CI: 21%-35%, k=18) for SPHOBIA; 9% (95% CI: 7%-11%, k=15) for social anxiety/social phobia; 13% (95% CI: 10%-16%, k=17) for GAD; 6% (95% CI: 4%-8%, k=14) for SAD; 9% (95% CI: 7%-12%, k=21) for OCD; 1% (95% CI: 0%-2%, k=4) for PD; 2% (95% CI: 0%-3%, k=3) for agoraphobia, and 2% (95% CI: 1%-3%, k=6) for PTSD (23).

Glasson et al. reported, in a sample of 909 people with 22q11.2 deletion syndrome, a prevalence range of oppositional defiant disorder of 10%-14%; mood disorder 3%-7%, and psychotic disorder 0%-5% (k=3). No CD was detected in a sample of 837 people with 22q11.2 deletion syndrome (k=2) (24).

### Prader Willi syndrome

No reviews of prevalence estimate of specific mental disorders in people with Prader-Willi syndrome were identified.

## Mental disorders in autistic people with ID

Although no meta-analyses specifically addressed the co-occurrence of ID and ASD, 8,791 autistic people with ID were included in six reviews primarily investigating people with ASD (27, 31, 33–35, 37).

Hollocks et al. conducted a subgroup analysis and reported a lower prevalence of *current depression* in autistic people with ID compared with those without ID [14% (95% CI: 5%-28%, N=512, k=6) vs 26% (95% CI: 20%-32%, N=1,413, k=16)] (27). In the same study, the prevalence of *any anxiety disorder* was 20% (95% CI: 7%-39%, N=394, k=6) in autistic people with ID versus 24% (95% CI: 19%-43%, N=1,050, k=7) in those without ID, while the prevalence of *OCD* was 24% (95% CI: 14%-36%, N=177, k=3) and 20% (95% CI: 10%-34%, N=970, k=7), respectively. The authors tested subgroup differences using meta-regression analyses, finding a significant difference for depression but not for anxiety or OCD (meta-regression coefficient for depression: β=0.12, p=0.03; not reported for anxiety and OCD) (27).

Regarding *psychosis,* Varcin et al. identified a prevalence of any psychotic disorder at 0%-32.6% (k=10) in 1,107 autistic adults with ID (34), and De Giorgi et al. reported non-affective psychosis prevalence range at 0%-25.1% (k=5) in 359 autistic people with ID across the lifespan (37). Curnow et al. reported a lifetime prevalence of psychosis of 10%-17% (k=2) in 4,208 autistic adolescents and adults with ID (35). Lugo-Marín et al. (33) reported prevalence estimates for schizophrenia spectrum disorders of 1.3%-26.1% (k=10) in autistic adults with ID.

*Mood disorders* ranged from 3.4%-36.1% (k=5) in autistic adults with ID (33), while Varcin et al. found rates of BD or manic episodes of 3.6%-6.2% (k=4) in 437 autistic adults with ID (34).

Curnow et al. reported a lifetime prevalence of *depression* of 13%-24% (k=2) in autistic adolescents and adults with ID (35).

Regarding *anxiety disorders*, Curnow et al. described a lifetime prevalence range of 17%-53%, and a current prevalence range of 40%-50% in autistic adolescents and adults with ID (k=2) (35). Higher estimates, at 41.7%-42.7% (k=3) for any anxiety or impairing anxiety level were shown by van Steensel et al. in autistic children and adolescents with ID (31), while Lugo-Marín et al. identified an anxiety disorder prevalence range of 0%-32.7% (k=7) in autistic adults with ID (33).

Regarding *specific anxiety disorders*, van Steensel et al. reported a prevalence range of GAD at 9.4%-24.6% (k=2), social anxiety disorder at 16.4%-19.9% (k=2), SAD at 10.5%-14.8% (k=2), and SPHOBIA at 17%-67.2% (k=3) in a population of 369 autistic children and adolescents with ID (31).

Lugo-Marín et al. reported a prevalence range for *personality disorder* at 0%-36.1% (k=6) and for SUD at 0%-11.9% (k=4) among autistic adults with ID (33).

## Mental disorders in offenders

Regarding offenders with ID, Rayner et al. described a prevalence range of 9.8%-80% (k=3) for any personality disorder, 22.1%-33% (k=2) for antisocial or borderline personality disorder, and 9.8% (k=2) for personality disorder not otherwise specified in a population of 372 offenders with ID (43). Torr et al. found a prevalence range for any personality disorder of 12%-54.8% (k=8), and 13%-38% (k=3) for antisocial personality disorder (44) in 769 offenders with ID.

### Mental disorders by setting

Regarding specific settings, Bakken et al. examined the characteristics and mental disorder diagnoses in 3,996 adults with ID admitted to inpatient units (32). The prevalence ranges reported for psychotic disorders ranged 6%-59% (k=19); for affective disorders (such as depression, mood disorder, BD, affective disorder, and mania) 9%-42% (k=19); for anxiety disorders (including anxiety and OCD) 0%-8%, excluding one outlier (k=18); and for other mental disorders (e.g., eating disorders, organic disorders, substance-related disorders, personality disorders, impulse control disorders, and dementias) showed prevalence rates ranging 0%-63% (k=17) (32).

Alexander et al. examined the published literature on the diagnosis of personality disorders in ID, reporting a prevalence range of <1%-91% in community settings, and between 22%-92% in hospital-based populations (k=14) (42).

## DISCUSSION

### Prevalence of mental disorders in people with intellectual disabilities

*Compared to the general population*, people with ID showed markedly higher prevalence rates for schizophrenia spectrum disorders (over six times higher), and OCD (nearly double, especially in children and adolescents). Anxiety disorders were also more frequent, though estimates varied by age. Compared to the general population, mood disorders appeared to be less prevalent, as were PTSD and personality disorders, possibly reflecting underdiagnosis (58, 59).

In *syndromic ID,* meta-analyses or subgroup analyses were available only for anxiety disorders, including subtypes such as SP, GAD, PD, PTSD, and OCD. Anxiety disorders were more prevalent in Fragile X syndrome, Williams syndrome, 22q11.2 deletion syndrome and less prevalent in Down syndrome.

Our umbrella review indicated that *ASD*, ADHD, and ID are often studied as separate conditions, with only one review conducting subgroup analysis comparing autistic adults with and without ID (27).

The small number of meta-analyses which stratified *data by ID severity or sex*, suggested higher rates of psychosis, OCD, anxiety, and depression in mild ID, and a slightly higher prevalence of psychosis in females than males.

### Findings in context

Several structural and methodological challenges hinder the accurate identification of mental disorders in people with ID. These include the limited availability of screening and diagnostic tools specifically designed or adapted for this population (60, 61), coupled with diagnostic overshadowing (62), whereby a mental disorder/symptom is attributed to the ID. Variability in the quality and accessibility of mental health services across countries (4, 63) also plays a role. For example, people with BIF often lack access to ID services, as eligibility criteria vary by region, hindering appropriate identification and care. These factors are likely to bias prevalence estimates, particularly in studies relying on clinical diagnoses rather than epidemiological investigations (64, 65).

Additional challenges arise from the debate of whether mental disorders may manifest as “behavioural equivalents”, or if such behavioural changes should never, or only rarely, be considered expressions of psychopathology (66, 67), especially in people with severe/profound ID and communication difficulties (68). This may help explain the higher reported rates of mental disorders in people with mild ID compared to those with more severe ID where diagnosis may not be possible to make.

Underdiagnosis remains largely unaddressed, as most studies and reviews did not consider bias from inconsistent application of diagnostic criteria. Although some reviews included primary studies using adapted criteria alongside DSM and/or ICD diagnoses, none reported prevalence data separately by diagnostic system versus adapted criteria, limiting the evaluation of their impact. Peña-Salazar et al. identified previously undiagnosed mental disorders in 29.6% of participants with ID, with major depressive and anxiety disorders being the most prevalent (69). Deb et al. similarly reported an increase in diagnosed mental disorders from 15.7% to 17.3% over 12 years in adults with ID in the Czech Republic, using routinely collected data (70).

Mood and depressive disorders illustrate how these challenges contribute to under-recognition or misdiagnosis in people with ID. Symptoms such as labile mood or irritability may be misinterpreted as behaviours that challenge or as psychosis. They may also go unrecognized due to a lack of adapted diagnostic criteria, or because people with ID may be unable to self-report core depressive features, including depressed mood or loss of interest and pleasure lasting at least two weeks (4, 60–63, 66–68). Our umbrella review reported a 2.8% prevalence estimate for depressive disorders in youth with ID (25) and 6.7% for any mood disorder (7), contrasting with recent primary studies (58, 59, 69) that suggest higher rates of depression. When adapted criteria are used, 13.7% may represent the lower limit of actual prevalence (58).

In this umbrella review, we included only studies reporting diagnoses of mental disorders, excluding those addressing mental health symptoms alone, as diagnoses offer greater specificity and comparability. While symptoms are observable indicators, they do not in themselves confirm a mental disorder (71). However, this is not the only approach to understanding mental health presentations in this population. For example, most of the literature on SUD, examined patterns of use and misuse alongside formal diagnoses (36, 39), illustrating the broad spectrum of substance-related behaviours seen in people with ID, which, though it captures clinically relevant phenomena, does not provide information about the number of people requiring some form of intervention.

## Strengths and limitations

This is the first umbrella review, to our knowledge, synthesising evidence across a wide range of mental disorders in diverse populations of people with ID (with or without co-occurring ASD) and allowing for comparison with prevalence data from the general population, where available. It also offers a comprehensive overview of the literature, including stratification by age, sex, level of ID, syndromic forms, and specific subgroups such as offenders or inpatients, thus enabling a broader understanding of comorbidity patterns in this population. However, umbrella reviews also have limitations. These include reliance on the methodological quality of the included reviews, the potential omission of primary studies not included in reviews, methodological difficulties in handling heterogeneity and overlapping data (72).

According to the AMSTAR-2 ratings, only one included review was rated as ’moderate’ quality (35), while the rest were rated ’low’ or ’critically low’. To support the correct interpretation of the results, we reported each original review’s AMSTAR-2 score (Table 1), provided confidence intervals when available, and included general population prevalence estimates for comparison.

Other limitations should be acknowledged. Definitions of ID, diagnostic criteria, and review methods have evolved, contributing to heterogeneity. For example, Mazza et al. used the category “mood disorder”, including both bipolar and depressive disorders (7), while diagnostic reclassifications (e.g., PTSD and OCD are not included in anxiety disorders in DSM-5) affected how some reviews grouped these conditions.

Variations in prevalence reporting timeframes (e.g., point, period, or lifetime prevalence) may limit the comparability of prevalence rates between the general population and people with ID. Most reviews included a higher proportion of males, which partly reflects the demographic distribution of people with ID. However, the potential for sex-specific differences has rarely been considered, limiting the generalisability and nuance of prevalence estimates.

Finally, our findings may also inform the debate on the relative risk contribution of ID for the onset of specific mental disorders. For example, our finding suggests that rates of depression in people with ID are lower than expected (58). However, investigating these associations was beyond the scope of our review, and caution is warranted when interpreting the findings, as the estimates were often based on few reviews or small samples, which restricts their generalisability.

### Clinical and Policy Implications

People with ID are at increased risk of developing comorbidities with mental disorders. Reliable, stratified prevalence estimates (e.g., by age, sex, or level of ID) can support accurate diagnosis and personalised care, guide service planning, inform the development of targeted interventions (e.g., for people with psychosis), and address currently unmet needs at individual, family and systems levels.

### Future research and gaps in the literature

High-quality systematic reviews and meta-analyses are lacking for several mental disorders in ID, including BD (and rapid cycling), eating disorders, SUD, and dementia, whether of syndromic or non-syndromic origin.

Evidence also remains limited for other disorders, such as PTSD, specific anxiety disorders, and personality disorders beyond antisocial/borderline types. Stratified prevalence data by sex, age, level of ID, and co-occurring neurodevelopmental conditions (e.g., ASD, ADHD, tic disorders) are still scarce. However, reviews by Lugo-Marín et al. and Varcin et al. identified multiple primary studies on various mental disorders in autistic people with ID, indicating that future meta-analyses may be achievable. Ecological studies are also needed to capture cultural and contextual variation of mental ill health (73). Finally, more clinical research is needed to improve diagnostic sensitivity and reduce underdiagnosis, together with validated tools adapted for people with ID (74), such as the SPAIDD-G (75), which, combined with clinical evaluation, shows promise in diagnosing mental disorders in people with ID.

Addressing these gaps is crucial to advancing our understanding of mental disorders in people with ID and improving their mental health care. Future research should prioritize high-quality systematic reviews and meta-analyses, alongside studies that explore underrepresented conditions and socio-demographic factors. By doing so, we can work towards more accurate prevalence estimates, culturally sensitive diagnostic frameworks, and tailored interventions that better meet the diverse needs of this population.

## Data Availability

Data availability is not applicable to this article as no new data were created or analysed in this study.

## Acknowledgements

The authors thank Dr. Chiara Ossola, Dr Mariasole Zanetti, and Dr Sofia Bosco for generously providing translation support on a voluntary basis. The authors would also like to thank Dr. Laura Colombo and Dr. Annalisa Bardelli from the Bibliographic Services of the University of Milano-Bicocca for their valuable support in developing and refining the search strategy. The authors thank Professor Muhammad Ayub for his inspiring insights, which helped shape the early ideas of this work. The authors made use of DeepL, ChatGPT, Google Translate to assist with the inclusion and assessment of not English primary studies. French and Spanish fluent speakers checked two full texts screenings and data retrieved in these languages.

## Author Contributions

H.A., B.C., B.G., S.J., B.M., and C.M. conceptualised the study. P.D., F.E., B.F., B.G., M.M., and S.J. curated the data. H.A., B.G., S.J., M.L., and M.M. performed the formal analysis. H.A., P.D., B.G., S.J., M.L., and M.M. contributed to the methodology. H.A. and S.J. supervised the project. B.G. and M.M. contributed to Writing-original draft. H.A., B.G., S.J., M.L., M.M., and R.S. contributed to Writing-review & editing. All authors contributed to data interpretation, critically revised the manuscript for important intellectual content, and approved the final version. B.G. and S.J. contributed equally to this work and are joint first authors.

