## Supplementary Materials for "Prevalence of mental disorders in people with intellectual disabilities across the lifespan: an umbrella review"

### Supplementary Table 1. SEARCH STRATEGY:

With the aim of providing a comprehensive overview of the prevalence of psychiatric disorders in individuals with intellectual disability, with or without co-occurring neurodevelopmental conditions (e.g., ASD), we updated the research protocol registered on PROSPERO (CRD42024610611). In the amended version (November 2024), we extended the age range to include the entire lifespan and reclassified references to ASD and ADHD from primary to secondary outcomes. This adjustment reflects our position that these conditions should not be considered psychiatric comorbidities, but rather distinct neurodevelopmental disorders.

With the assistance of the bibliographic services of the University of Milano-Bicocca the following MeSh Terms and filters were adapted and searched through seven databases: the Cochrane Library, MEDLINE, PubMed, PsycINFO (ProQuest), Embase, CINAHL and Epistemonikos. 
To point out that for 4 databases (ProQuest, Cochrane, CINAHL, and Epistemonikos) the operator and MeSh Term "AND Comorbidity" was not used due to the limited number of reports found. 
 
**PUBMED:** 

| MeSh Terms | (Developmental Disabilities OR Intellectual Disability OR Persons with Mental Disabilities OR Learning Disabilities OR Specific Learning Disorder) AND (Comorbidity OR comorbid OR comorbidities OR comorbids) |
| --- | --- |
| String | (("developmental disabilities"[MeSH Terms] OR ("developmental"[All Fields] AND "disabilities"[All Fields]) OR "developmental disabilities"[All Fields] OR ("intellectual disability"[MeSH Terms] OR ("intellectual"[All Fields] AND "disability"[All Fields]) OR "intellectual disability"[All Fields]) OR ("persons with mental disabilities"[MeSH Terms] OR ("persons"[All Fields] AND "mental"[All Fields] AND "disabilities"[All Fields]) OR "persons with mental disabilities"[All Fields]) OR ("learning disabilities"[MeSH Terms] OR ("learning"[All Fields] AND "disabilities"[All Fields]) OR "learning disabilities"[All Fields]) OR ("specific learning disorder"[MeSH Terms] OR ("specific"[All Fields] AND "learning"[All Fields] AND "disorder"[All Fields]) OR "specific learning disorder"[All Fields])) AND ("comorbid"[All Fields] OR "comorbidity"[MeSH Terms] OR "comorbidity"[All Fields] OR "comorbidities"[All Fields] OR "comorbids"[All Fields] OR ("comorbid"[All Fields] OR "comorbidity"[MeSH Terms] OR "comorbidity"[All Fields] OR "comorbidities"[All Fields] OR "comorbids"[All Fields]) OR ("comorbid"[All Fields] OR "comorbidity"[MeSH Terms] OR "comorbidity"[All Fields] OR "comorbidities"[All Fields] OR "comorbids"[All Fields]) OR ("comorbid"[All Fields] OR "comorbidity"[MeSH Terms] OR "comorbidity"[All Fields] OR "comorbidities"[All Fields] OR "comorbids"[All Fields]))) AND ((meta-analysis[Filter] OR systematicreview[Filter])) |
| Filters applied | meta-analysis or systematic review |

### AMSTAR TOOL EVALUATION AND ASSESSMENT:

#### Supplementary Table 2. Notes for scoring

| **N** | **ITEM** | **Considerations for scoring**  i**n this umbrella review** |
| --- | --- | --- |
| **1** | Did the research questions and inclusion criteria for the review include the components of PICO? | Comparison**:** the presence or absence of a comparison group is not consistent with our scope; it was therefore not penalised in the scoring. |
| **2** | Did the report of the review contain an explicit statement that the review methods were established prior to conduct of the review and did the report justify any significant deviations from the protocol? | Considered not applicable if the systematic review was published before the introduction of the PROSPERO platform in February 2011. |
| **3** | Did the review authors explain their selection of the study designs for inclusion in the review? |  |
| **4** | Did the review authors use a comprehensive literature search strategy? |  |
| **5** | Did the review authors perform study selection in duplicate? |  |
| **6** | Did the review authors perform data extraction in duplicate? |  |
| **7** | Did the review authors provide a list of excluded studies and justify the exclusions? |  |
| **8** | Did the review authors describe the included studies in adequate detail? |  |
| **9** | Did the review authors use a satisfactory technique for assessing the risk of bias  (RoB) in individual studies that were included in the review? | Only validated tools or clear risk-of-bias descriptions were considered satisfactory techniques for RoB. |
| **10** | Did the review authors report on the sources of funding for the studies included in the review? |  |
| **11** | If meta-analysis was justified did the review authors use appropriate methods for statistical combination of results? | Appropriate methods for statistical combination of results have been restricted to the presence of heterogeneity tests (i.e. I²) and sensitivity analyses, as other analysis may not have been performed due to insufficient data. |
| **12** | If meta-analysis was performed did the review authors assess the potential impact  of RoB in individual studies on the results of the meta-analysis or other evidence synthesis? |  |
| **13** | Did the review authors account for RoB in individual studies when interpreting/ discussing the results of the review? |  |
| **14** | Did the review authors provide a satisfactory explanation for, and discussion of, any heterogeneity observed in the results of the review? |  |
| **15** | If they performed quantitative synthesis did the review authors carry out an adequate investigation of publication bias (small study bias) and discuss its likely impact on the results of the review? | For some meta-analyses, it was not possible to conduct a quantitative analysis of heterogeneity due to the inclusion of too few primary studies. Full points were awarded if heterogeneity was addressed in the discussion and/or results section, especially when the authors discussed their findings in relation to other populations, as in Royston et al. |
| **16** | Did the review authors report any potential sources of conflict of interest, including any funding they received for conducting the review? |  |

Critical domains: 2, 4, 7, 9, 11, 13, 15

#### Supplementary Table 3. Amstar 2 critical domains and rating

**AMSTAR 2 critical domains**

• Protocol registered before commencement of the review (item 2)

• Adequacy of the literature search (item 4)

• Justification for excluding individual studies (item 7)

• Risk of bias from individual studies being included in the review (item 9)

• Appropriateness of meta-analytical methods (item 11)

• Consideration of risk of bias when interpreting the results of the review (item 13)

• Assessment of presence and likely impact of publication bias (item 15)

*From: Shea B J, Reeves B C, Wells G, Thuku M, Hamel C, Moran J et al. AMSTAR 2: a critical appraisal tool for systematic reviews that include randomised or non-randomised studies of healthcare interventions, or both BMJ 2017; 358 :j4008 doi:10.1136/bmj.j4008*

| **Rating overall confidence in the results of the review** | |
| --- | --- |
| **High** | No or one non-critical weakness: the systematic review provides an accurate and comprehensive summary of the results of the available studies that address the question of interest |
| **Moderate** | More than one non-critical weakness*: the systematic review has more than one weakness but no critical flaws. It may provide an accurate summary of the results of the available studies that were included in the review |
| **Low** | One critical flaw with or without non-critical weaknesses: the review has a critical flaw and may not provide an accurate and comprehensive summary of the available studies that address the question of interest |
| **Critical Low** | More than one critical flaw with or without non-critical weaknesses: the review has more than one critical flaw and should not be relied on to provide an accurate and comprehensive summary of the available studies |

***1*** *From: Shea B J, Reeves B C, Wells G, Thuku M, Hamel C, Moran J et al. AMSTAR 2: a critical appraisal tool for systematic reviews that include randomised or non-randomised studies of healthcare interventions, or both BMJ 2017; 358:j4008 doi:10.1136/bmj.j4008.* 
**Multiple non-critical weaknesses may diminish confidence in the review, and it may be appropriate to move the overall appraisal down from moderate to low confidence.*

#### Supplementary Table 4. AMSTAR-2: item by item scoring

| **Amstar -2 ITEM** | **Alexander 2003** | **Aman 2016** | **Bakken 2013** | **Buckley 2020** | **Curnow**  **2023** | **Daveney 2019** | **De Giorgi 2019** | **Edwards 2022** | **Glasson**  **2020** | **Holloks**  **2019** | **Huxley 2019** | **Lugo-Marín 2019** | **Maïano 2018** |
| --- | --- | --- | --- | --- | --- | --- | --- | --- | --- | --- | --- | --- | --- |
| 1 | No | Yes | Yes | Yes | Yes | Yes | Yes | Yes | Yes | Yes | Yes | Yes | Yes |
| 2 | NA | No | No | Yes | Yes | No | No | Yes | No | No | No | Yes | No |
| 3 | No | Yes | Yes | Yes | Yes | Yes | Yes | Yes | Yes | yes | Yes | Yes | Yes |
| 4 | No | PY | PY | PY | Yes | PY | PY | PY | PY | PY | No | PY | PY |
| 5 | No | Yes | No | Yes | Yes | No | Yes | No | Yes | Yes | Yes | Yes | Yes |
| 6 | No | No | No | Yes | No | No | Yes | No | Yes | No | Yes | Yes | Yes |
| 7 | No | No | No | No | PY | No | Yes | No | No | No | No | No | PY |
| 8 | PY | Yes | Yes | PY | Yes | Yes | Yes | Yes | Yes | Yes | PY | Yes | Yes |
| 9 | No | Yes | No | Yes | Yes | Yes | Yes | Yes | Yes | Yes | PY | Yes | Yes |
| 10 | No | No | No | No | No | Yes | No | No | No | no | No | No | No |
| 11 | NoMa | Yes | NoMa | Yes | NoMa | Yes | NoMa | Yes | Yes | Yes | NoMa | Yes | Yes |
| 12 | NoMa | Yes | NoMa | Yes | NoMa | Yes | NoMa | Yes | Yes | yes | NoMa | Yes | Yes |
| 13 | No | Yes | No | Yes | Yes | Yes | Yes | Yes | Yes | No | Yes | Yes | Yes |
| 14 | No | Yes | No | Yes | Yes | Yes | Yes | Yes | Yes | yes | Yes | Yes | Yes |
| 15 | NoMa | Yes | NoMa | Yes | NoMa | Yes | NoMa | Yes | Yes | yes | NoMa | Yes | Yes |
| 16 | Yes | Yes | No | yes | Yes | Yes | Yes | Yes | Yes | yes | Yes | Yes | Yes |
| **Critical weaknesses** | 4 | 2 | 4 | 1 | 0 | 2 | 1 | 1 | 2 | 2 | 3 | 1 | 1 |
| **Non-critical weaknesses** | 6 | 2 | 5 | 1 | 2 | 2 | 1 | 3 | 1 | 3 | 1 | 1 | 1 |
| **Quality Evaluation** | **CRITICAL LOW** | **CRITICAL LOW** | **CRITICAL LOW** | **LOW** | **MODERATE** | **CRITICAL LOW** | **LOW** | **LOW** | **CRITICAL LOW** | **CRITICAL LOW** | **CRITICAL LOW** | **LOW** | **LOW** |

| **Amstar -2 ITEM** | **Mazza et al 2020** | **Mevissen 2010** | **Oeseburg**  **2011** | **Rayner**  **2015** | **Reardon 2015** | **Royston**  **2017** | **Torr**  **2008** | **van Duijvenbode 2019** | **van Steensel**  **2011** | **Varcin 2022** | **Walton**  **2015** | **Walton**  **2016** | **Whitaker**  **2006** |
| --- | --- | --- | --- | --- | --- | --- | --- | --- | --- | --- | --- | --- | --- |
| 1 | Yes | Yes | Yes | Yes | Yes | Yes | Yes | Yes | Yes | Yes | Yes | Yes | Yes |
| 2 | Yes | NA | NA | No | No | No | NA | No | NA | Yes | No | No | NA |
| 3 | Yes | No | Yes | Yes | Yes | Yes | No | Yes | Yes | Yes | Yes | Yes | Yes |
| 4 | PY | No | PY | PY | PY | PY | No | PY | PY | PY | Yes | PY | PY |
| 5 | Yes | No | Yes | No | No | No | No | No | No | Yes | No | Yes | No |
| 6 | Yes | No | No | No | No | No | No | No | No | Yes | no | No | No |
| 7 | No | No | No | No | No | PY | No | PY | No | No | No | No | No |
| 8 | Yes | PY | Yes | PY | Yes | Yes | Yes | PY | Yes | Yes | Yes | Yes | Yes |
| 9 | Yes | No | Yes | No | No | Yes | No | No | Yes | PY | Yes | Yes | No |
| 10 | No | No | No | No | No | No | No | No | No | No | No | No | No |
| 11 | Yes | NoMa | NoMa | NoMa | NoMa | Yes | NoMa | NoMa | Yes | Yes | NoMa | NoMa | NoMa |
| 12 | Yes | NoMa | NoMa | NoMa | NoMa | Yes | NoMa | NoMa | Yes | Yes | NoMa | NoMa | NoMa |
| 13 | Yes | No | No | No | Yes | Yes | No | No | Yes | Yes | Yes | Yes | No |
| 14 | Yes | Yes | Yes | Yes | No | No | no | Yes | Yes | Yes | Yes | Yes | Yes |
| 15 | Yes | NoMa | NoMa | NoMa | NoMa | Yes | NoMa | NoMa | Yes | Yes | NoMa | NoMa | NoMa |
| 16 | Yes | No | No | No | No | Yes | No | Yes | No | Yes | No | No | No |
| **Critical weaknesses** | 1 | 4 | 2 | 4 | 3 | 1 | 4 | 3 | 1 | 1 | 2 | 2 | 3 |
| **Non-critical weaknesses** | 1 | 5 | 3 | 4 | 5 | 4 | 6 | 3 | 4 | 1 | 4 | 3 | 4 |
| **Quality Evaluation** | **LOW** | **CRITICAL LOW** | **CRITICAL LOW** | **CRITICAL LOW** | **CRITICAL LOW** | **LOW** | **CRITICAL LOW** | **CRITICAL LOW** | **LOW** | **LOW** | **CRITICAL LOW** | **CRITICAL LOW** | **CRITICAL LOW** |

**Table 4.** *NA*: Not applicable; *PY*: Partially yes; *NoMa*: No meta-analysis conducted.

### List of studies included/excluded at full-text screening

#### Studies identified through database searching and excluded

| **N°** | **Autor’s Names, Year** | **Title, scientific journal, Vol, Issue, DOI** | **Evaluation** |
| --- | --- | --- | --- |
| 1. | Abregú-Cresp R, et.al, (2024) | School bullying in children and adolescents with neurodevelopmental and psychiatric conditions: a systematic review and meta-analysis. The Lancet Child & Adolescent Health, 8(2): 122 - 134 | Population not within the scope of this review and report out of remit. It focuses on the broader neurodevelopmental population and on school bullying, which is not a recognised psychiatric diagnosis. |
| 2. | Adams D, et al. (2023) | Longitudinal studies of challenging behaviours in autistic children and adults: A systematic review and meta-analysis. Clinical Psychology Review 104:102320 | Population not within the scope of this review and report out of remit. It focuses on individuals with ASD, and challenging behaviours are not considered psychiatric diagnostic categories according to the DSM. |
| 3. | Akker, N., 2021 | Behavioural, psychiatric and psychosocial factors associated with aggressive behaviour in adults with intellectual disabilities: A systematic review and narrative analysis. Journal of Applied Research in Intellectual Disabilities Vol. 34 Issue 2 Pages 327-389 | Population not within the scope of this review and report out of remit. It focuses on factors associated with aggressive behaviour in the ADHD population, no prevalence data on psychiatric comorbidities in the ID population were retrieved. |
| 4. | Aman, M. G., et al. (2004) | Treatment of behavior disorders in mental retardation: report on transitioning to atypical antipsychotics, with an emphasis on risperidone. The Journal of clinical psychiatry 65(9): 1197-1210. | Not a systematic review and report out of remit. It is a report which focuses on transitioning to atypical antipsychotics. |
| 5. | Aman LCS, et al. (2024) | Psychotic illness in people with Prader-Willi syndrome: a systematic review of clinical presentation, course and phenomenology. Orphanet J Rare Dis. 2024 Feb 15;19(1):69. doi: 10.1186/s13023-024-03026-y. PMID: 38360662; PMCID: PMC10870655. | Report out of remit. It focuses on the prevalence of psychotic illness based on specific genetic subtypes in individuals with Prader–Willi syndrome. |
| 6. | Andersson, A., et al. (2020). | Research Review: The strength of the genetic overlap between ADHD and other psychiatric symptoms - a systematic review and meta-analysis. Journal of child psychology and psychiatry, and allied disciplines 61(11): 1173-1183. | Population not within the scope of this review and report out of remit. It focuses on genetic overlap between people with ASD and psychiatric symptoms, no prevalence data. |
| 7. | Aymerich C, et al., (2024) | Prevalence and Correlates of the Concurrence of Autism Spectrum Disorder and Obsessive-Compulsive Disorder in Children and Adolescents: A Systematic Review and Meta-Analysis. Brain Sci., 14(4), 379 | Population not within the scope of this review. It focuses on people with ASD and no prevalence data of interest were retrieved for the ID population. |
| 8. | Ayub, M., et al. (2015) | Clozapine for psychotic disorders in adults with intellectual disabilities. Cochrane Database of Systematic Reviews 9(9): CD010625. | Report out of remit. It focuses on medication, no prevalence data on psychiatric disorders. |
| 9. | Bakken, T. L. (2021) | Behavioural equivalents of schizophrenia in people with intellectual disability and autism spectrum disorder. A selective review. International Journal of Developmental Disabilities 67(5): 310-317. | Report out of remit. It focuses on behavioural equivalents, no prevalence data on psychiatric disorders. |
| 10. | Bennett, C. (2018) | Prescribing psychotropic medication to people with intellectual disability. Australian and New Zealand Journal of Psychiatry 52(1): 76. | Report out of remit. It focuses on prescribing medication, no prevalence data on psychiatric disorders. |
| 11. | Blickwedel, J., et al. (2019) | Epilepsy and challenging behaviour in adults with intellectual disability: A systematic review. Journal of Intellectual and Developmental Disability 44(2): 219-231. | Report out of remit. It focuses on epilepsy and challenging behaviours, no prevalence data on the psychiatric comorbidities of interest in the ID population. |
| 12. | Boon, L. C., et al. (2017) | Catatonia in patients with autism spectrum disorder (asd): Comorbidity, risk factors, and treatment. Developmental Medicine and Child Neurology 59: 105. | Population not within the scope of this review.  It focuses on ASD population. |
| 13. | Bora, E. and R. M. Murray (2014) | Meta-analysis of cognitive deficits in ultra-high risk to psychosis and first-episode Psychosis: Do the cognitive deficits progress over, or after, the onset of psychosis? Schizophrenia Bulletin 40(4): 744-755. | Report out of remit. It focuses on cognitive deficits, no prevalence data on psychiatric disorders. |
| 14. | Bourne, J., et al. (2022) | A systematic review of community psychosocial group interventions for adults with intellectual disabilities and mental health conditions. Journal of Applied Research in Intellectual Disabilities 35(1): 3-23. | Report out of remit. It focuses on interventions, no prevalence data on psychiatric disorders. |
| 15. | Brosnan, J. and O. Healy (2011) | A review of behavioral interventions for the treatment of aggression in individuals with developmental disabilities. Research in developmental disabilities 32(2): 437-446. | Population not within the scope of this review and report out of remit. It focuses on interventions for anger and aggression in people with developmental disabilities, no prevalence data on psychiatric disorders. |
| 16. | Browne, C. and I. C. Smith (2018) | Psychological interventions for anger and aggression in people with intellectual disabilities in forensic services. Aggression & Violent Behavior 39: 1-14. | Report out of remit. It focuses on interventions for anger and aggression, no prevalence data on psychiatric disorders. |
| 17. | Bruinsma, E., et al. (2020) | Non‐pharmacological interventions for challenging behaviours of adults with intellectual disabilities: A meta‐analysis. Journal of Intellectual Disability Research 64(8): 561-578. | Report out of remit. It focuses on interventions for challenging behaviours, no prevalence data. |
| 18. | Brylewski, J. and L. Duggan (1999) | Antipsychotic medication for challenging behaviour in people with intellectual disability: a systematic review of randomized controlled trials. Journal of intellectual disability research: JIDR 43 ( Pt 5)(5): 360-371. | Report out of remit. It focuses on medication for challenging behaviours, no prevalence data. |
| 19. | Buckles, J., et al. (2013). | A systematic review of the prevalence of psychiatric disorders in adults with intellectual disability, 2003–2010. Journal of Mental Health Research in Intellectual Disabilities 6(3): 181-207 | Report out of remit. It focuses primarily on general prevalence rates of any psychiatric comorbidity and symptoms, rather than on specific disorders of interest. |
| 20. | Bundock, K. E. and O. Hewitt (2017). | A review of social skills interventions for adults with autism and intellectual disability. Tizard Learning Disability Review 22(3): 148-158. | Report out of remit. It focuses on interventions, no prevalence data. |
| 21. | Byrne, G. (2020) | A Systematic Review of Treatment Interventions for Individuals with Intellectual Disability and Trauma Symptoms: A Review of the Recent Literature. Trauma, violence & abuse: 1524838020960219. | Report out of remit. It focuses on treatment, no prevalence data. |
| 22. | Carroll Chapman SL, Wu LT. (2012) | Substance abuse among individuals with intellectual disabilities. Res Dev Disabil. 2012 Jul-Aug;33(4):1147-56. doi: 10.1016/j.ridd.2012.02.009. Epub 2012 Mar 7. PMID: 22502840; PMCID: PMC3328139. | Report out of remit. It aims to identify common findings and knowledge gaps concerning substance use problems. Excluded due to significant heterogeneity and lack of a clear diagnostic definition of substance misuse. |
| 23. | Cassidy, S. A., et al. (2018) | Measurement properties of tools used to assess depression in adults with and without autism spectrum conditions: A systematic review. Autism Res 11(5): 738-754 | Report out of remit. It focuses on ACEs, no prevalence data. |
| 24. | Chaplin, R. (2004) | General psychiatric services for adults with intellectual disability and mental illness. Journal of intellectual disability research: JIDR 48(1): 1-10. | Report out of remit. It aims to assess differences in outcome for patients with ID and mental disorders treated in general or specialised ID mental health services. |
| 25. | Chaplin, R. (2009) | Annotation. New research into general psychiatric services for adults with intellectual disability and mental illness. Journal of Intellectual Disability Research 53(3): 189-199. | Report out of remit. It aims to assess differences in outcome for patients with ID and mental disorders treated in general or specialised ID mental health services |
| 26. | Colijn MA, et al. (2024) | SETD1A variant-associated psychosis: A systematic review of the clinical literature and description of two new cases. Progress in Neuro-Psychopharmacology and Biological Psychiatry, 8. | Report out of remit. It aims to characterize the neurodevelopmental and psychiatric phenotypes of SETD1A variant-associated schizophrenia |
| 27. | Cooper, N. C., Balachandran Nair, D., Egan, S., Barrie, A., & Perera, B. (2019). | Current evidence for the identification and management of premenstrual syndrome in women with intellectual disabilities: A systematic review. Advances in Mental Health and Intellectual Disabilities, 13(6), 268–283. | Not a systematic review. It is an expert review of the evidence on PMD in ID. Regarding prevalence it says that 1 study PMS has significantly higher prevalence in autism than a matched control population (11% vs 92%, po 0.0000001). |
| 28. | Courtial E, et al. (2024) | Antipsychotic medication in people with intellectual disability and schizophrenia: A 25-year updated systematic review and cross-sectional study. Journal of Psychopharmacology;38(12):1045-1053 | Report out of remit. It focuses on diagnostic overshadowing. No prevalence data on psychiatric comorbidities in individuals with Intellectual disability were retrieved. |
| 29. | Crompton, C., et al. (2021) | A systematic review of adverse childhood experiences (ACEs) with people with intellectual disabilities: an unsafe gap in the literature. Advances in Mental Health & Intellectual Disabilities 15(5): 158-170 | Report out of remit. It focuses on ACEs, no prevalence data. |
| 30. | Dagnan, D., et al. (2018). | A systematic review of cognitive behavioural therapy for anxiety in adults with intellectual disabilities. J Intellect Disabil Res 62(11): 974-991. | Report out of remit. It focuses on CBT, no prevalence data. |
| 31. | Danielsson, H., Imms, C., Ivarsson, M. et al. (2024) | A Systematic Review of Longitudinal Trajectories of Mental Health Problems in Children with Neurodevelopmental Disabilities. J Dev Phys Disabil 36, 203–242. https://doi.org/10.1007/s10882-023-09914-8 | Population not within the scope of this review. It focuses on neurodevelopmental disorders (NDD). Although some of the included primary studies address Fragile X syndrome, intellectual disability, or developmental delay, the review itself aims to highlight trends in mental health outcomes measured using psychometric scales. This focus does not align with the primary objective of our review, as it does not report any prevalence data. |
| 32. | Danquah, A., et al. (2009) | An investigation of factors predictive of continued self-injurious behaviour in an intellectual disability service. Journal of Applied Research in Intellectual Disabilities 22(4): 395-399. | Report out of remit. It focuses on self-injurious behaviour predictive factors, no prevalence data on specific psychiatric comorbidities |
| 33. | Davies, B., et al. (2015) | The relationship between emotional recognition ability and challenging behaviour in adults with an intellectual disability: A systematic review. Journal of Intellectual Disabilities 19(4): 393-406 | Report out of remit. It focuses between emotional recognition ability and challenging behaviour, no prevalence data. |
| 34. | Day C et al (2016) | Intellectual disability and substance use/misuse: a narrative review", Journal of Intellectual Disabilities and Offending Behaviour, Vol. 7 Iss 1 pp. 25 - 34 Permanent link to this document: http://dx.doi.org/10.1108/JIDOB-10-2015-0041 | Not a systematic review. It provides a general overview of prevalence, risk factors, and interventions, but does not present original data. |
| 35. | de Winter, C. F., et al. (2011) | Physical conditions and challenging behaviour in people with intellectual disability: A systematic review. Journal of Intellectual Disability Research 55(7): 675-698. | Report out of remit. It focuses on physical conditions and challenging behaviour. No prevalence data on the psychiatric comorbidities of interest in the ID population. |
| 36. | Deb, S., et al. (2008) | The effectiveness of mood stabilizers and antiepileptic medication for the management of behaviour problems in adults with intellectual disability: A systematic review. Journal of Intellectual Disability Research 52(2): 107-113. | Report out of remit. It focuses on medication, no prevalence data. |
| 37. | Deb, S., et al. (2014) | The effectiveness of aripiprazole in the management of problem behaviour in people with intellectual disabilities, developmental disabilities and/or autistic spectrum disorder--a systematic review. Research in Developmental Disabilities 35(3): 711-725. | Report out of remit. It focuses on medication, no prevalence data. |
| 38. | Deb, S., et al. (2020) | Association between epilepsy and challenging behaviour in adults with intellectual disabilities: Systematic review and meta-analysis. BJPsych Open 6. | Report out of remit. It focuses on association between epilepsy and challenging behaviours, no prevalence data on specific psychiatric comorbidities. |
| 39. | Deb, S., et al. (2022) | The European guideline on the assessment and diagnosis of psychiatric disorders in adults with intellectual disabilities. European Journal of Psychiatry 36(1): 11-25. | Report out of remit and not a systematic review. It is a guideline for assessment and diagnosis of psychiatric disorders, no prevalence data. |
| 40. | Dell’Armo K and Tassé MJ, (2024) | Diagnostic Overshadowing of Psychological Disorders in People With Intellectual Disability: A Systematic Review. Am J Intellect Dev Disabil, 129 (2): 116–134 | Report out of remit. It focuses on diagnostic overshadowing. No prevalence data on psychiatric comorbidities in individuals with Intellectual disability were retrieved. |
| 41. | den Brok, W. L. and Sterkenburg P.S. (2015) | Self-controlled technologies to support skill attainment in persons with an autism spectrum disorder and/or an intellectual disability: a systematic literature review. Disability and rehabilitation. Assistive technology 10(1): 1-10. | Report out of remit. It focuses on self-controlled technologies, no prevalence data. |
| 42. | Dhamija, D, et al. (2023) | Evaluation of Efficacy of Cannabis Use in Patients With Attention Deficit Hyperactivity Disorder: A Systematic Review. Cureus,15(6):e40969 | Population not within the scope of this review and report out of remit. It focuses on understanding the nature of the relationship between cannabis use and ADHD symptoms. |
| 43. | Dhiman V et al., (2023) | A systematic review and meta-analysis of prevalence of seven psychiatric disorders in India. Indian Journal Psychiatry; 65(11):1096–1103 | Report out of remit. It describes the percentage of ID and others disorders in India. |
| 44. | Di Luzio M, et al., (2023) | Clinical features and comorbidity in very early-onset schizophrenia: a systematic review. Frontiers in Psychiatry,13;14:1270799. | Report out of remit. It focuses on the clinical features of very early-onset schizophrenia, and no prevalence data on individuals with intellectual disability were retrieved. |
| 45. | Dodd, P., et al. (2005) | A review of the emotional, psychiatric and behavioural responses to bereavement in people with intellectual disabilities. Journal of Intellectual Disability Research 49(7): 537-543. | Report out of remit. It focuses on responses to bereavement, on the psychiatric comorbidities of interest in this ID population. |
| 46. | Dodd P, Doherty A, Guerin S. (2016) | A Systematic Review of Suicidality in People with Intellectual Disabilities. Harv Rev Psychiatry. 2016 May-Jun;24(3):202-13. doi: 10.1097/HRP.0000000000000095. PMID: 27148912. | Report out of remit. It focuses on identifying the nature of research on suicidality in individuals with intellectual disability and includes one primary study reporting incidence data on suicidality in this population. |
| 47. | Eaton, C., et al. (2021) | A systematic review of the behaviours associated with depression in people with severe–profound intellectual disability. Journal of Intellectual Disability Research 65(3): 211-229. | Report out of remit. It focuses on behaviours, no prevalence data. |
| 48. | Einfeld, S. L., et al. (2011) | Comorbidity of intellectual disability and mental disorder in children and adolescents: a systematic review. J Intellect Dev Disabil 36(2): 137-143. | Report out of remit. It focuses on estimating the rates of mental disorder in the ID population, no prevalence data on specific psychiatric comorbidities. |
| 49. | Ellena Wood, Neel Halder  (2014) | Gender disorders in learning disability – a systematic review TIZARD LEARNING DISABILITY REVIEW j VOL. 19 NO. 4 2014, pp. 158-165, C Emerald Group Publishing Limited, ISSN 1359-5474 | Population not within the scope of this review and not a systematic review. It addresses the entire learning disability population with a broad focus on aetiology, treatment, and management. Additionally, gender dysphoria is not considered a psychiatric disorder by experts in the field, which further limits its relevance to our aims. |
| 50. | Espinet SD, et al. (2022) | A Review of Canadian Diagnosed ADHD Prevalence and Incidence Estimates Published in the Past Decade. Brain Science, 12(8), 1051 | Population not within the scope of this review. It focuses on individuals with ADHD. |
| 51. | Fahrendorff AM, et al. (2023) | Psychiatric comorbidity in patients with pediatric bipolar disorder – A systematic review. Acta Psychiatrica Scandinavica.148:110–132 | Population not within the scope of this review. It focuses on people with pediatric bipolar disorder, and no data on intellectual disability were collected. |
| 52. | Fazel, S., et al. (2008) | The prevalence of intellectual disabilities among 12000 prisoners--A systematic review. International Journal of Law and Psychiatry 31(4): 369-373. | Report out of remit. It focuses on the prevalence of intellectual disabilities in prison populations, no prevalence data on the psychiatric comorbidities of interest in this ID population. |
| 53. | Ferreira B.R., Pio-Abreu J.L., Januario C. | Tourette’s syndrome and associated disorders: A systematic review. Trends in Psychiatry and Psychotherapy 2014 36:3 (123-133) | Population not within the scope of this review and report out of remit. It focuses on Tourette’s syndrome, and overall ID prevalence. Non pooled data with other psychiatric comorbidities. |
| 54. | Flavell J, Franklin C, Nestor PJ. (2022) | A Systematic Review of Fragile X-Associated Neuropsychiatric Disorders. J Neuropsychiatry Clin Neurosci. 2023 Spring;35(2):110-120. doi: 10.1176/appi.neuropsych.21110282. Epub 2022 Sep 29. PMID: 36172690. | Population not within scope of this review and report out of remit. It focuses in reviewing both fragile X premutation prevalence in patients with neurodevelopmental disorders and psychiatric disorder prevalence in premutation carriers without fragile X-associated tremor/ataxia syndrome (FXTAS). |
| 55. | Flynn, S., et al. (2017) | Measurement tools for mental health problems and mental well-being in people with severe or profound intellectual disabilities: A systematic review. Clinical Psychology Review 57: 32-44. | Report out of remit. It focuses on measurements tool, no prevalence data. |
| 56. | Forde J., et al (2022) | Health Status of Adults with Autism Spectrum Disorder. Review Journal Autism and Developmental Disorders 9, 427–437 | Population not within the scope of this review. It focuses on individuals with ASD. No prevalence data on psychiatric comorbidities in individuals with Intellectual disability were retrieved. |
| 57. | Francés L., et al. (2022) | Psychiatry and Mental Current state of knowledge on the prevalence of neurodevelopmental disorders in childhood according to the DSM-5: a systematic review in accordance with the PRISMA criteria. Child and Adolescent Health, 16, 27 | Report out of remit. It aims to establish the prevalence all neurodevelopmental disorders globally. |
| 58. | Froude AM, et al. (2024) | The prevalence of cannabis use disorder in attention-deficit hyperactivity disorder: A clinical epidemiological meta-analysis. Journal of Psychiatric Research, 172, 391-401 | The population is not within the scope of this review, as it focuses on individuals with ADHD. |
| 59. | Froude AM, et al. 2024 | The prevalence of cannabis use disorder in attention-deficit hyperactivity disorder: A clinical epidemiological meta-analysis, Journal of Psychiatric Research, Volume 172, 2024, Pages 391-401, ISSN 0022-3956, https://doi.org/10.1016/j.jpsychires.2024.02.050. | Population not within the scope of this review and report out of remit. It focuses on ADHD population. No prevalence data on ID or reference to IQ. |
| 60. | Furniss, F. and A. B. Biswas (2012) | Recent research on aetiology, development and phenomenology of self‐injurious behaviour in people with intellectual disabilities: A systematic review and implications for treatment. Journal of Intellectual Disability Research 56(5): 453-475. | Report out of remit. It focuses on self‐injurious behaviour, no prevalence data on the psychiatric comorbidities of interest in the ID population. |
| 61. | Gerhand S. & Saville CWN, (2022) | ADHD prevalence in the psychiatric population. International Journal of Psychiatry in Clinical Practice, 26, Issue 2 | Population not within the scope of this review. It focuses on people with ADHD. No prevalence data on psychiatric comorbidities in individuals with intellectual disability were retrieved. |
| 62. | Gilderthorp, R. C. (2015) | Is EMDR an effective treatment for people diagnosed with both intellectual disability and post-traumatic stress disorder? Journal of intellectual disabilities : JOID 19(1): 58-68. | Report out of remit. It focuses on the EMDR treatment for ID people with PTSD, no prevalence data. |
| 63. | Gormez, A., et al. (2014) | Pharmacological interventions for self-injurious behaviour in adults with intellectual disabilities: Abridged republication of a Cochrane systematic review. Journal of psychopharmacology (Oxford, England) 28(7): 624-632. | Report out of remit. It focuses on the treatment of self-injurious behaviours, no prevalence data. |
| 64. | Gregori, E., et al. (2018) | Treatment of Self-Injurious Behavior in Adults with Intellectual and Developmental Disabilities: A Systematic Review. Journal of Developmental & Physical Disabilities 30(1): 111-139. | Report out of remit. It focuses on the treatment of self-injurious behaviours, no prevalence data. |
| 65. | Gustafsson, C., et al. (2009) | Effects of psychosocial interventions for people with intellectual disabilities and mental health problems: A survey of systematic reviews. Research on Social Work Practice 19(3): 281-290. | Report out of remit. It focuses on interventions, no prevalence data. |
| 66. | Gyereh, J & Shukla M. (2023) | Risk factors of and interventions for mental health problems in learning disabilities: A systematic review of psychological therapies for parents and children. Current Psychology, 43(5):1-17 | Report out of remit. It focuses on psychological therapies for parents and children. |
| 67. | Haasbroek H & Morojele N, (2022) | A Systematic Literature Review on the Relationship Between Autism Spectrum Disorder and Substance Use Among Adults and Adolescents. Review Journal of Autism and Developmental Disorders, 9, 1–20 | Population not within scope. It focuses on people with ASD. No prevalence data in the ID population retrieved. |
| 68. | Halvorsen MB, et al., (2023) | General Measurement Tools for Assessing Mental Health Problems Among Children and Adolescents with an Intellectual Disability: A Systematic Review. Journal of Autism and Developmental Disorders. 53(1):132-204. | Report out of remit. It focuses on psychometric properties of tools assessing general mental health problems in children (4–20 yrs) with ID. No prevalence data on psychiatric diagnoses in individuals with Intellectual Disabilities were retrieved. |
| 69. | Hamelin, J., et al. (2013) | Anger management and intellectual disabilities: A systematic review. Journal of Mental Health Research in Intellectual Disabilities 6(1): 60-70 | Report out of remit. It focuses on anger management, no prevalence data. |
| 70. | Hamers, P. C. M., et al. (2018) | Non‐pharmacological interventions for adults with intellectual disabilities and depression: A systematic review. Journal of Intellectual Disability Research 62(8): 684-700. | Report out of remit. It focuses on interventions, no prevalence data. |
| 71. | Hammond, S. and N. Beail (2020) | The relationship between cognitive variables and offending behaviour in adults with intellectual disabilities: A systematic review. Journal of Applied Research in Intellectual Disabilities 33(4): 779-792. | Report out of remit. It focuses on interventions for aggressive behaviours, no prevalence data. |
| 72. | Harris EC, Barraclough B. (1997) | Suicide as an outcome for mental disorders. A meta-analysis. Br J Psychiatry. 1997 Mar;170:205-28. doi: 10.1192/bjp.170.3.205. PMID: 9229027. | Report out of remit. Prevalence data for the intellectual disability population are presented as odds ratio, which does not align with our focus on prevalence estimates. |
| 73. | Hassiotis, A. A., et al. (2015) | Behavioural and cognitive-behavioural interventions for outwardly-directed aggressive behaviour in people with intellectual disabilities. Cochrane Database of Systematic Reviews(4)  DOI: 10.1002/14651858.CD003406.pub4 | Report out of remit. It focuses on interventions for aggressive behaviours, no prevalence data. |
| 74. | Hellenbach, M., et al. (2017) | Intellectual disabilities among prisoners: Prevalence and mental and physical health comorbidities. Journal of Applied Research in Intellectual Disabilities 30(2): 230-241. | Report out of remit. No prevalence data on the psychiatric comorbidities of interest in the ID population found. It focuses on describing the prevalence of ID among prisoners, and physical and psychiatric comorbidities (in terms of associations). |
| 75. | Hermans, H. and H. M. Evenhuis (2010) | Characteristics of instruments screening for depression in adults with intellectual disabilities: Systematic review. Research in Developmental Disabilities 31(6): 1109-1120. | Report out of remit. It focuses on instruments screening for depression, no prevalence data. |
| 76. | Hermans, H., et al. (2011) | Instruments assessing anxiety in adults with intellectual disabilities: A systematic review. Research in Developmental Disabilities 32(3): 861-870. | Report out of remit. It focuses on assessment of anxiety, no prevalence data. |
| 77. | Heyvaert, M., et al. (2014) | Systematic review of restraint interventions for challenging behaviour among persons with intellectual disabilities: Focus on effectiveness in single‐case experiments. Journal of Applied Research in Intellectual Disabilities 27(6): 493-510. | Report out of remit. It focuses on restrain interventions for challenging behaviours, no prevalence data. |
| 78. | Hounsome, J., et al. (2018) | The structured assessment of violence risk in adults with intellectual disability: A systematic review. Journal of Applied Research in Intellectual Disabilities 31(1): e1-e17 | Report out of remit. It focuses on assessment of violence risk, no prevalence data. |
| 79. | Irazabal Gimenez, M. (2016) | The family burden of caregivers of young and adult people diagnosed with intellectual disability and mental disorders: A systematic review. Psiquiatria Biologica 23(3): 93-102. | Report out of remit. It focuses on family burden of caregivers, no prevalence data. |
| 80. | Jeevanandam L. (2009) | Perspectives of intellectual disability in Asia: epidemiology, policy, and services for children and adults. Curr Opin Psychiatry. 2009 Sep;22(5):462-8. doi: 10.1097/YCO.0b013e32832ec056. PMID: 19625968. | Not a systematic review. It is an expert umbrella review. |
| 81. | Joseph, J. K., & Devu, B. K. (2022). | Prevalence and Pattern of Learning Disability in India: A Systematic Review and Meta-Analysis. JAMA Oncol, (3):420–444. | Population not within scope of this review. It focuses on people with learning disabilities. |
| 82. | Kerr, S., et al. (2013) | Tobacco and alcohol-related interventions for people with mild/moderate intellectual disabilities: a systematic review of the literature. Journal of intellectual disability research: JIDR 57(5): 393-408. | Report out of remit. It focuses on interventions of interventions, no prevalence data. |
| 83. | Khoury E, et al. (2023) | Meta-analysis of personal and familial co-occurrence of Attention Deficit/Hyperactivity Disorder and Bipolar Disorder. Neuroscience & Biobehavioral Reviews,146, 105050 | Population not within scope of this review. It focuses on people with ADHD. No prevalence data were retrieved for people with Intellectual Disabilities |
| 84. | Kildahl, A. N., Bakken, T. L., Iversen, T. E., & Helverschou, S. B. (2019). | Identification of Post-Traumatic Stress Disorder in Individuals with Autism Spectrum Disorder and Intellectual Disability: A Systematic Review. *Journal of Mental Health Research in Intellectual Disabilities*, *12*(1–2), 1–25. https://doi.org/10.1080/19315864.2019.1595233 | Review out of remit. It explores the presentation of PTSD in autistic people with intellectual disabilities and aims to identify knowledge gaps in its recognition and diagnosis within this population. No relevant prevalence data are provided. |
| 85. | Kwetsie H, van Schaijk M, Van Der Lee S, Maes-Festen D, Ten Hoopen LW, van Haelst MM, Coesmans M, Van Den Berg E, De Wit MCY, Pijnenburg Y, Aronica E, Boot E, Van Eeghen AM. (2024) | Dementia in Rare Genetic Neurodevelopmental Disorders: A Systematic Literature Review. Neurology. 2024 Jun 11;102(11):e209413. doi: 10.1212/WNL.0000000000209413. Epub 2024 May 17. PMID: 38759134; PMCID: PMC11175636. | Report out of remit. It provides a systematic overview of current knowledge on dementia, cognitive and adaptive trajectories, and associated factors in adults with rare genetic neurodevelopmental disorders (RGNDs). Three primary studies reported prevalence data separately for three different syndromic forms of intellectual disability. |
| 86. | Kim JH, et al. (2024) | Association of self-harm and suicidality with psychiatric co-occurring conditions in autistic individuals  eClinicalMedicine,77: 102863 | Population not within scope of this review and report out of remit. It focuses on self-harm, and suicide (not a psychiatric comorbidity) in people with ASD. No incidence data were retrieved for people with Intellectual Disabilities. |
| 87. | Koslowski, N., et al. (2016) | Effectiveness of interventions for adults with mild to moderate intellectual disabilities and mental health problems: systematic review and meta-analysis. Br J Psychiatry 209(6): 469-474. | Report out of remit. It focuses on interventions, no prevalence data. |
| 88. | Kumar S, Lata S, Verma S and Anupriya, (2024) | Associated Factors of Suicidal Behavior Among Persons with Physical Disability: A Systematic Review. Indian Journal of Psychological Medicine, 11;46(4):298–304 | Population not within scope of this review. It focuses on people with general physical disability. |
| 89. | Kuo, S. S. and S. M. Eack (2020) | Meta-analysis of cognitive performance in neurodevelopmental disorders during adulthood: Comparisons between autism spectrum disorder and schizophrenia on the Wechsler Adult Intelligence Scales. Frontiers in Psychiatry 11 | Report out of remit. It focuses on cognitive performance, no prevalence data. |
| 90. | La Malfa, G., et al. (2006) | Reviewing the use of antipsychotic drugs in people with intellectual disability. Human Psychopharmacology: Clinical and Experimental 21(2): 73-89. | Report out of remit. It focuses on antipsychotic medications, no prevalence data. |
| 91. | Lee J.y., Patel M. & Scior K., (2023) | Self-esteem and its relationship with depression and anxiety in adults with intellectual disabilities: a systematic literature review, Journal of Intellectual Disability Research, 67, 6 | Report out of remit. It focuses more on mechanism, and no prevalence data were retrieved for psychiatric comorbidities in people with Intellectual Disabilities |
| 92. | Lees-Warley, G. and Rose J. (2015) | What does the evidence tell us about adults with low intellectual functioning who deliberately set fires? A systematic review. International Journal of Developmental Disabilities 61(4): 242-256. | Report out of remit. It focuses on psychosocial characteristics of firesetters and the outcomes of treatment interventions, no prevalence data on the psychiatric comorbidities of interest in the ID population. |
| 93. | Lofthouse, R., et al. (2017) | How effective are risk assessments/measures for predicting future aggressive behaviour in adults with intellectual disabilities (ID): A systematic review and meta-analysis. Clinical Psychology Review 58: 76-85. | Report out of remit. It focuses on risk assessment and measures for prediction, no prevalence data. |
| 94. | Melville, C. A., et al. (2016) | Statistical modelling studies examining the dimensional structure of psychopathology experienced by adults with intellectual disabilities: Systematic review." Research in Developmental Disabilities 53-54: 1-10 | Report out of remit. It focuses on statistical modelling studies examining the dimensional structure of psychopathology, no prevalence data. |
| 95. | Metcalfe, D., et al. (2020) | Screening tools for autism spectrum disorder, used with people with an intellectual disability: A systematic review. Res. Autism Spectr. Disord. 74. | Report out of remit. It focuses on screening tools for ASD, no prevalence data. |
| 96. | Micai M, et al. (2023) | Prevalence of co-occurring conditions in children and adults with autism spectrum disorder: A systematic review and meta-analysis. Neuroscience & Biobehavioral Reviews, 155, 105436 | Population not within scope. It focuses on people with ASD, and no prevalence data were retrieved for people with both ASD and Intellectual Disability regarding psychiatric comorbidities. |
| 97. | Mingins JE, Tarver J, Waite J, Jones C, Surtees AD. (2021) | Anxiety and intellectual functioning in autistic children: A systematic review and meta-analysis. Autism. 2021 Jan;25(1):18-32. doi: 10.1177/1362361320953253. Epub 2020 Nov 16. PMID: 33198481; PMCID: PMC8162138. | Population not within the scope of this review. It focuses con ASD, and reports on anxiety symptoms combined with anxiety disorders. It uses correlations for statistics between ID and non ID groups. |
| 98. | Mishra, S., et al., (2024) | Prevalence of adult attention deficit hyperactivity disorder in India: a systematic review and a cross-sectional study among young adults in Delhi-NCR. Social Psychiatry and Psychiatric Epidemiology | Population not within scope. It focuses on people with ADHD. |
| 99. | Molina-Ruiz, R. M., et al. (2017) | A guide to psychopharmacological treatment of patients with intellectual disability in psychiatry. Int J Psychiatry Med 52(2): 176-189. | Report out of remit. It’s a selective review on psychopharmacological treatment. |
| 100. | Mollison, E., et al. (2014) | A review of risk factors associated with suicide in adults with intellectual disability. Advances in Mental Health & Intellectual Disabilities 8(5): 302-308 | Report out of remit. It focuses on risk factors, no incidence data. |
| 101. | Morinaga, M., et al. (2020) | Migration or ethnic minority status and risk of autism spectrum disorders and intellectual disability: systematic review." European journal of public health  DOI: 10.1093/eurpub/ckaa108 | Report out of remit. It focuses on the relation between ASD or ID and migration or ethnic minority status, no prevalence data |
| 102. | Murray A., et al. (2022) | Autism, Problematic Internet Use and Gaming Disorder: A Systematic Review, Comprehensive Psychiatry, 118,152346 | Population not within scope of this review. It focuses on people with ASD. |
| 103. | Mutluer T, et al. (2022) | Population-Based Psychiatric Comorbidity in Children and Adolescents With Autism Spectrum Disorder: A Meta-Analysis, Frontiers of Psychiatry, 1. | Population not within scope of this review. It focuses on individuals with ASD. No prevalence data were retrieved for individuals with both ASD and Intellectual Disability regarding psychiatric comorbidities. |
| 104. | N Buckley1, E Glasson1, W Chen2, A Epstein1, H Leonard1, R Skoss1, M Blackmore3, J Downs1 | The prevalence and phenotypic signatures of mental ill heath across neurogenetic disorders associated with intellectual disability: A systematic review | Not a systematic review. It is a conference paper |
| 105. | Nagdee M . (2011) | Dementia in intellectual disability: a review of diagnostic challenges. Afr J Psychiatry (Johannesbg). 2011 Jul;14(3):194-9. doi: 10.4314/ajpsy.v14i3.1. PMID: 21863203. | Not a systematic review. It is an expert review of dementia in ID. |
| 106. | Nagdee, M. (2011) | Dementia in intellectual disability: a review of diagnostic challenges." Afr J Psychiatry (Johannesbg) 14(3): 194-199. | Report out of remit. It focuses on dementia-related diagnostic challenges, no prevalence data on the psychiatric comorbidities of interest in the ID population. |
| 107. | Nicoll, M., et al. (2013) | Cognitive Behavioural Treatment for Anger in Adults with Intellectual Disabilities: A Systematic Review and Meta-analysis. Journal of Applied Research in Intellectual Disabilities 26(1): 47-62. | Report out of remit. It focuses on treatment for anger, no prevalence data. |
| 108. | Normand CL, et al. (2022) | A Systematic Review of Problematic Internet Use in Children, Adolescents, and Adults with Autism Spectrum Disorder, Review Journal of Autism and Developmental Disorders, 9, 507–520 | Population not within scope of this umbrella review. It focuses on people with ASD. |
| 109. | O’Halloran L, et al. (2022) | Suicidality in autistic youth: A systematic review and meta-analysis, Clinical Psychology Review, 93,102144 | Population not within scope of this resview and report out of remit. It focuses on people with ASD. |
| 110. | Oliphant RYK, Smith EM, Grahame V. | What is the Prevalence of Self-harming and Suicidal Behaviour in Under 18s with ASD, With or Without an Intellectual Disability? J Autism Dev Disord. 2020 Oct;50(10):3510-3524. doi: 10.1007/s10803-020-04422-6. PMID: 32125568. | Report out of remit. It focuses on suicidal ideation, suicide attempts, suicidal behavior (including thoughts, plans, and actions), self-harm, and combined suicidal behaviors in the ASD population. The influence of intellectual disability (ID) on prevalence is reported only as the percentage of participants who "often or very often" talk about death or suicide, which falls outside the remit of our umbrella review. |
| 111. | Osugo, M. and S. A. Cooper (2016) | Interventions for adults with mild intellectual disabilities and mental ill‐health: A systematic review." Journal of Intellectual Disability Research 60(6): 615-622. | Report out of remit. It focuses on interventions, no prevalence data. |
| 112. | Padgett, F. E., et al. (2010) | The co-occurrence of nonaffective psychosis and the pervasive developmental disorders: A systematic review." Journal of Intellectual and Developmental Disability 35(3): 187-198. | Population not within the scope of this review, study out of remit. It focuses on pervasive developmental disorders, no prevalence data on non-affective psychosis in the ID population |
| 113. | Pedapati, E. V. (2019) | 3.3 Assessment and treatment of common psychiatric concerns in individuals with intellectual disability disorder." Journal of the American Academy of Child and Adolescent Psychiatry 58(10): S134. | Report out of remit. It focuses on assessment and treatment, no prevalence data. |
| 114. | Peltopuro M, Ahonen T, Kaartinen J, Seppälä H, Närhi V. (2014) | Borderline intellectual functioning: a systematic literature review. Intellect Dev Disabil. 2014 Dec;52(6):419-43. doi: 10.1352/1934-9556-52.6.419. PMID: 25409130. | Not a systematic review. It is an expert overview of intellectual functioning. It addresses various aspects including neurocognitive functioning, social behaviours, mental health (with references to four primary studies investigating mental health problems in individuals with BIF), employment and marriage, as well as risk and protective factors. |
| 115. | Peltopuro, M., et al. (2014) | Borderline intellectual functioning: A systematic literature review. Intellectual and Developmental Disabilities 52(6): 419-443 | Population not within the scope of this review, and out of remit. It focuses on borderline intellectual functioning and highlights that adult with BIF face various challenges and socioeconomic disadvantages and reported risk/protective factors are not BIF-specific. No prevalence data on the psychiatric comorbidities of interest in the ID population. |
| 116. | Peña-Salazar, C., Arrufat, F., Santos, J. M., Novell, R., & Valdés-Stauber, J. (2018). | Psychopathology in borderline intellectual functioning: A narrative review. Advances in Mental Health and Intellectual Disabilities, 12(1), 22–33. [https://doi.org/10.1108/AMHID-07-2017-0031](https://psycnet.apa.org/doi/10.1108/AMHID-07-2017-0031) | Report out of remit. The systematic review does not report absolute prevalence rates and presents data only in terms of odds ratios or indirect comparisons. Therefore, it is not aligned with our focus on quantitative prevalence estimates. |
| 117. | Perez-Achiaga, N., et al. (2009) | Instruments for the detection of depressive symptoms in people with intellectual disabilities: a systematic review." Journal of Intellectual Disabilities 13(1): 55-76. | Report out of remit. It focuses on instruments for depressive symptoms detection, no prevalence data. |
| 118. | Priday, L. J., et al. (2017) | Behavioural interventions for sleep problems in people with an intellectual disability: a systematic review and meta-analysis of single case and group studies." Journal of intellectual disability research : JIDR 61(1): 1-15. | Report out of remit. It focuses on interventions for sleep problems, no prevalence data. |
| 119. | Pridding, A. and N. G. Procter (2008) | A systematic review of personality disorder amongst people with intellectual disability with implications for the mental health nurse practitioner." Journal of Clinical Nursing 17(21): 2811-2819 | Not a systematic review. This is an umbrella review. |
| 120. | Pridding, A., et al. (2007) | Mental health nursing roles and functions in acute inpatient units: caring for people with intellectual disability and mental health problems--a literature review. The international journal of psychiatric nursing research 12(2): 1459-1471. | Report out of remit. It focuses on mental health nursing roles and functions no prevalence data. |
| 121. | Pruijssers, A. C., et al. (2014) | The relationship between challenging behaviour and anxiety in adults with intellectual disabilities: a literature review. Journal of Intellectual Disability Research 58(2): 162-171 | Report out of remit. It focuses on the relationship between challenging behaviour and anxiety, no prevalence data on the psychiatric comorbidities of interest. |
| 122. | Purper-Ouakil D., and Weibel S. (2025) | Comorbidite´ s et diagnostics diffe´ rentiels du Trouble De´ficit de l’Attention Hyperactivite´ (TDAH) en fonction de l’aˆge, Ann Med Psychol (Paris), <https://doi.org/10.1016/j.amp.2024.09.006> | Population not within scope of this review, and not a systematic review. It is an expert review focusing on the ADHD population. Reviewed by two French-speaking clinicians; no prevalence data on the population with intellectual disability were reported. |
| 123. | Rana, F., et al. (2013) | Pharmacological interventions for self‐injurious behaviour in adults with intellectual disabilities. Cochrane Database of Systematic Reviews(4).  DOI: 10.1002/14651858.CD009084.pub2 | Report out of remit. It focuses on interventions, no prevalence data. |
| 124. | Roy, A., et al. (2015) | Are opioid antagonists effective in reducing self-injury in adults with intellectual disability? A systematic review. Journal of Intellectual Disability Research 59(1): 55-67. | Report out of remit. It focuses on opioid antagonists effectiveness in reducing self-injury, no prevalence data on the psychiatric comorbidities of interest. |
| 125. | Royle, A. (2013). | Alcohol and illicit substance misuse in individuals with intellectual disabilities, University of Oxford (UK), 2013 - 157 | Not a systematic review and report out of remit. It is a thesis which focuses on treatment and interventions for illicit drugs and alcohol misuse and investigates the attitudes of healthcare staff working in ID services to substance misuse and treatment intervention. |
| 126. | Russell PSS et al, (2022) | Prevalence of intellectual disability in India: A meta-analysis. World Journal of Clinal Pediatrics,9;11(2) | Report out of remit. It focuses on establishing the summary prevalence of intellectual disability during the past 60 years in India, and not the presence of psychiatric comorbidity in the ID population. |
| 127. | Sainsbury WJ, et al. (2022) | Age of Diagnosis for Cooccurring Autism and Attention Deficit Hyperactivity Disorder During Childhood and Adolescence: a Systematic Review, International Journal of Developmental Neuroscience, 82,8 | Population not within scope of this review. It focuses on ASD, and ADHD. No prevalence data retrieved for the population with intellectual disability |
| 128. | Sawyer, A., et al. (2014) | Psychopharmacological treatment of challenging behaviours in adults with autism and intellectual disabilities: A systematic review. Research in Autism Spectrum Disorders 8(7): 803-813. | Report out of remit. It focuses on psychopharmacological treatment of challenging behaviours, no prevalence data on the psychiatric comorbidities of interest. |
| 129. | Schmid R., Picarel F., Campbell R., Bougeard C.,Buitelaar J. | P.0048 Prevalence of co-morbidities in autistic children and adolescents in five european countries and the united states: a systematic literature review. [European Neuropsychopharmacology](https://www.sciencedirect.com/journal/european-neuropsychopharmacology)  [Volume 53, Supplement 1](https://www.sciencedirect.com/journal/european-neuropsychopharmacology/vol/53/suppl/S1), December 2021, Pages S35-S36 | Not a systematic review, it is a conference abstract. |
| 130. | Shanahan, P. J., et al. (2019) | Interventions for sleep difficulties in adults with an intellectual disability: a systematic review. J Intellect Disabil Res 63(5): 372-385. | Report out of remit. It focuses on interventions for sleep disorders, no prevalence data on the psychiatric comorbidities of interest. |
| 131. | Sharma, E., Sharma, L. P., Balachander, S., Lin, B., Manohar, H., Khanna, P., Lu, C., Garg, K., Thomas, T. L., Au, A. C. L., Selles, R. R., Højgaard, D. R. M. A., Skarphedinsson, G., & Stewart, S. E. (2021). | Comorbidities in Obsessive-Compulsive Disorder Across the Lifespan: A Systematic Review and Meta-Analysis. Frontiers in psychiatry, 12, 703701. https://doi.org/10.3389/fpsyt.2021.703701 | Report out of remit. It focuses on OCD. Neurodevelopmental disorders considered separately, and no combined data for people with ID retrieved. |
| 132. | Sheehan, R. and A. Hassiotis (2017) | Reduction or discontinuation of antipsychotics for challenging behaviour in adults with intellectual disability: a systematic review. The Lancet. Psychiatry 4(3): 238-256. | Report out of remit. It focuses on antipsychotics discontinuation for challenging behaviours, no prevalence data on the psychiatric comorbidities of interest. |
| 133. | Sheerin, F., et al. (2019) | Medication management in intellectual disability settings: A systematic review. Journal of intellectual disabilities : JOID: 1744629519886184. | Report out of remit. It focuses on medication management, no prevalence data on the psychiatric comorbidities of interest. |
| 134. | Simpson, M. K. and J. Hogg (2001). | Patterns of offending among people with intellectual disability: A systematic review. Part I: Methodology and prevalence data." Journal of Intellectual Disability Research 45(5): 384-396. | Report out of remit. It focuses on patterns of offending, no prevalence data on the psychiatric comorbidities of interest. |
| 135. | Simpson, M. K. and J. Hogg (2001). | Patterns of offending among people with intellectual disability: a systematic review. Part II: predisposing factors. Journal of Intellectual Disability Research 45(5): 397-406 | Report out of remit. It focuses on patterns of offending, no prevalence data on the psychiatric comorbidities of interest. |
| 136. | Smit, M. J., et al. (2019) | Clinical characteristics of individuals with intellectual disability who have experienced sexual abuse. An overview of the literature. Research in developmental disabilities 95: 103513. | Report out of remit. It focuses on clinical characteristics of people with ID who experienced sexual abuse. No prevalence data on the psychiatric comorbidities of interest. |
| 137. | Sohanpal, S. K., et al. (2007) | The effectiveness of antidepressant medication in the management of behaviour problems in adults with intellectual disabilities: A systematic review. Journal of Intellectual Disability Research 51(10): 750-765. | Report out of remit. It focuses on the effectiveness of antidepressant medication in the management of challenging behaviours. No prevalence data on the psychiatric comorbidities of interest. |
| 138. | St John, L., et al. (2020) | A Systematic Review and Meta-Analysis Examining the Effect of Exercise on Individuals With Intellectual Disability." American journal on intellectual and developmental disabilities 125(4): 274-286 | Report out of remit. It focuses on the effect of exercise, no prevalence data on the psychiatric comorbidities of interest. |
| 139. | Takagi S, et al. (2022) | Motor Functional Characteristics in Attention-Deficit/Hyperactivity Disorder and Autism Spectrum Disorders: A Systematic Review, Neuropsychiatric Disease and Treatment, 18, 1679–1695 | Population not within scope of this review and report out of remit. It focuses on ADHD and ASD population and no prevalence data retrieved for the population with intellectual disability. |
| 140. | Thomson, A., et al. (2009) | Amfetamine for attention deficit hyperactivity disorder in people with intellectual disabilities. Cochrane Database of Systematic Reviews(1)  DOI: 10.1002/14651858.CD007009.pub2 | Population not within the scope of this review and report out of remit. It focuses on medication in the ADHD population, no prevalence data. |
| 141. | Thomson, A., et al. (2009) | Risperidone for attention‐deficit hyperactivity disorder in people with intellectual disabilities. Cochrane Database of Systematic Reviews(2)  DOI: 10.1002/14651858.CD007011.pub2 | Population not within the scope of this review and report out of remit. It focuses on medication in the ADHD population. |
| 142. | Tomsa, R., et al. (2021) | Prevalence of Sexual Abuse in Adults with Intellectual Disability: Systematic Review and Meta-Analysis. International journal of environmental research and public health 18(4): 1-17. | Report out of remit of this review. It focuses on sexual abuse, no prevalence data on the psychiatric comorbidities of interest. |
| 143. | Tromans, S., et al. (2018) | The Prevalence of intellectual disability  s in Adult Psychiatric Inpatients: A Systematic Review. Clinical practice and epidemiology in mental health : CP & EMH 14: 177-187 | Population not within the scope of this review. It focuses on people with ASD. |
| 144. | van de Wouw, E., et al. (2012) | Prevalence, associated factors and treatment of sleep problems in adults with intellectual disability: a systematic review. Research in developmental disabilities 33(4): 1310-1332. | Report out of remit. It focuses on associated factors and treatment of sleep problems, without providing prevalence data on the psychiatric comorbidities of interest. |
| 145. | van Ool JS, et al (2016) | A systematic review of neuropsychiatric comorbidities in patients with both epilepsy and intellectual disability. Epilepsy Behav. 2016 Jul;60:130-137. doi: 10.1016/j.yebeh.2016.04.018. Epub 2016 May 18. PMID: 27206231. | Report out of remit. It aims to identify which neuropsychiatric comorbidities are typical in individuals with both epilepsy and intellectual disability, and to explore the factors associated with these comorbidities. No prevalence data are reported. |
| 146. | Vanstraelen M, Tyrer SP (1999). | Rapid cycling bipolar affective disorder in people with intellectual disability: a systematic review. J Intellect Disabil Res. 1999 Oct;43 ( Pt 5):349-59. doi: 10.1046/j.1365-2788.1999.043005349.x. PMID: 10546959. | Report out of remit. It focuses on systematically reviewing case studies and small series involving patients with a dual diagnosis of rapid-cycling bipolar affective disorder and intellectual disability, providing detailed information on individual mood cycles, including the length and frequency of episodes over a one-year period. No prevalence data relevant to our scope are provided. |
| 147. | Vaquerizo-Serrano J, Salazar De Pablo G, Singh J, Santosh P. (2022) | Catatonia in autism spectrum disorders: A systematic review and meta-analysis. Eur Psychiatry. 2021 Dec 15;65(1):e4. doi: 10.1192/j.eurpsy.2021.2259. PMID: 34906264; PMCID: PMC8792870. | Report out of remit. It provides evidence of catatonic features in individuals with autism spectrum disorder. Only one primary study included a sample with more than 50% of participants diagnosed with intellectual disability. |
| 148. | Verdugo, M. A., et al. (2020). | A systematic review of the assessment of support needs in people with intellectual and developmental disabilities. Int. J. Environ. Res. Public Health 17(24): 1-26. | Report out of remit. It focuses on the assessment of support needs, no prevalence data. |
| 149. | Virués-Ortega, J., et al. (2014) | Clinical decision making and preference assessment for individuals with intellectual and developmental disabilities. American Journal on Intellectual and Developmental Disabilities 119(2): 151-170. | Report out of remit. It focuses on decisional aspects and assessment, no prevalence data. |
| 150. | Welch, K. A., et al. (2011) | Systematic review of the clinical presentation of schizophrenia in intellectual disability. Journal of Psychopathology and Behavioral Assessment 33(2): 246-253. | Report out of remit. It focuses on clinical aspects of schizophrenia, no prevalence data retrieved. |
| 151. | Werling AM, et al. (2022) | Problematic use of digital media in children and adolescents with a diagnosis of attention-deficit/ hyperactivity disorder compared to controls. A meta-analysis, Journal of Behavioral Addictions, 11, 2, 305–325 | Population not within scope of this review and report out of remit. It focuses on ADHD population and no prevalence data retrieved for the population with intellectual disability. |
| 152. | Werner, S. and M. Stawski (2012) | Mental health: knowledge, attitudes and training of professionals on dual diagnosis of intellectual disability and psychiatric disorder. Journal of intellectual disability research : JIDR 56(3): 291-304. | Report out of remit. It focuses examining the knowledge, attitudes and training of psychiatrists and other professional caregivers in regard to serving people with DD, not prevalence. |
| 153. | Weyrauch D, Schwartz M, Hart B, Klug MG, Burd L. (2017) | Comorbid Mental Disorders in Fetal Alcohol Spectrum Disorders: A Systematic Review. J Dev Behav Pediatr. 2017 May;38(4):283-291. doi: 10.1097/DBP.0000000000000440. PMID: 28460370. | Population not within the scope of this review and report out of remit. It focuses on FAS and overall ID prevalence is not combined with other psychiatric comorbidities. |
| 154. | White, M. J., et al. (1995). | Diagnostic overshadowing and mental retardation: A meta-analysis. American Journal on Mental Retardation 100(3): 293-298. | Report out of remit. It focuses on diagnostic overshadowing and, not prevalence. |
| 155. | Williams, E. M. and J. Rose (2020) | Nonpharmacological treatment for individuals with intellectual disability and "personality disorder". J Appl Res Intellect Disabil 33(4): 767-778. | Report out of remit. It focuses on non-pharmacological treatment, not prevalence. |
| 156. | Zeilinger, E. L., et al. (2013) | A systematic review on assessment instruments for dementia in persons with intellectual disabilities. Research in developmental disabilities 34(11): 3962-3977. | Report out of remit. It focuses on assessment instrument, not prevalence. |
| 157. | Zeilinger, E. L., et al. (2013). | CAPs- IDD: Characteristics of Assessment Instruments for Psychiatric Disorders in Persons with Intellectual Developmental Disorders. Journal of Intellectual Disability Research 57(8): 737-746. | Report out of remit. It focuses on assessment instrument, not prevalence. |
| 158. | Zeilinger, E. L., et al. (2020) | Informant-based assessment instruments for dementia and their measurement properties in persons with intellectual disability: systematic review protocol. BMJ open 10(12): e040920. | Report out of remit. It focuses on assessment instrument and measurement, not prevalence. |
| 159. | Zeilinger, E. L., et al. (2021) | Informant-based assessment instruments for dementia in people with intellectual disability: A systematic review and standardised evaluation." Research in developmental disabilities 121: 104148. | Report out of remit. It focuses on assessment instrument, not prevalence. |

#### Supplementary Table 6. Studies identified through manual searching and excluded

| N* | **Authors, Year** | **Title, Journal, Vol, Iss** | **Reason for the exclusion** |
| --- | --- | --- | --- |
| 1 | Azam, K., Sinai, A., Hassiotis, A. 2009 | Mental ill-health in adults with learning disabilities, Psychiatry, Volume 8, Issue 10, October 2009, Pages 376-381 | Not a systematic review. It is an expert review of epidemiological studies on mental health in people with ID. No search strategy retrieved. |
| 2 | Bougeard C, et al. 2021 | Prevalence of Autism Spectrum Disorder and Co-morbidities in Children and Adolescents: A Systematic Literature, Front Psychiatry. Oct 27;12:744709. | Population not within scope of this review and report out of remit. The focus is on the prevalence of ASD and co-morbidities in children and adolescents. No prevalence data on psychiatric comorbidity in the ID population retrieved. |
| 3 | Bowring, Darren L.^a,;^ Painter, Jon^c^; Hastings, Richard P.^a, d,^ 2019 | Prevalence of Challenging Behaviour in Adults with Intellectual Disabilities, Correlates, and Association with Mental Health, Current Developmental Disorders Reports 6(2):1-9 | Report out of remit. It focuses on challenging behaviours, not on psychiatric comorbidities. |
| 4 | Brown, C.M., Newell, V., Sahin, E., Hedley, D.2024 | Updated Systematic Review of Suicide in Autism: 2018–2024, Curr Dev Disord Rep **11**, 225–256 | Population not within scope of this review and report out of remit. It focuses on suicide in ASD; no prevalence/incidence data on the ID population. |
| 5 | Carr, A., O'Reilly, G.2016 | Diagnosis, classification and epidemiology, In: A. Carr, C. Linehan, G. O' Reilly, P. Walsh, & J. McEvoy (eds). Handbook of Intellectual Disability and Clinical Psychology Practice (Second Edition). London: Routledge. , pp.3-44 | Not a systematic review. It is is a book chapter. |
| 6 | Chan, W.M.R., Bhandarkar, R.2024 | Suicidality and Intellectual Disability: A Systematic Review, Journal of Mental Health Research in Intellectual Disabilities Volume 18, 2025 - Issue 1 | Report out of remit. It focuses primarily on the prevalence (with no data to establish a precise estimate), risk factors, and interventions for suicidality among individuals with intellectual disability. |
| 7 | Cooper, S.-A., Bertelli, M.O., Bradley, E.2022 | Epidemiology of Psychiatric Disorders in Persons with Intellectual Disabilities. In: Bertelli, M.O., Deb, S.(., Munir, K., Hassiotis, A., Salvador-Carulla, L. (eds) Textbook of Psychiatry for Intellectual Disability and Autism Spectrum Disorder. Springer, Cham. https://doi.org/10.1007/978-3-319-95720-3_9 | Not a systematic review. It is a book chapter. |
| 8 | Cooper, Van Der Speck, Rohan, 2009 | Epidemiology of mental ill health in adults with intellectual disabilities, Curr Opin Psychiatry. Sep;22(5):431-6. | Not a systematic review. It is not reported a defined search strategy. It is an expert review of mental health epidemiological studies relevant to adults with intellectual disabilities, published from January 2008 to early 2009. |
| 9 | EAST W.N.1932 | Mental defectiveness and alcohol and drug addiction, Brit. J. Inebr. 29: 149-168  1932 | Not retrieved. |
| 10 | Escamilla-Soto MC, Montoya-Rojas YA, Quintero-Cadavid CP, García-Valencia J. (2024) | Trastornos depresivos y ansiosos en población con discapacidad intelectual. Iatreia [Internet]. 6 de mayo de 2024 [citado 6 de mayo de 2025];37(4). Disponible en: https://revistas.udea.edu.co/index.php/iatreia/article/view/350203 | Not a systematic review; limited search (PubMed and Google Scholar only) |
| 11 | Fledderman, N., Clemente, E., Merrick, J., Patel, D.R.2022 | Intellectual disability (intellectual developmental disorder). In Nova Science Publishers, Inc, Int J Child Health Hum Dev 2022;15(3):225-235 | Not a systematic review. It’s a book chapter. |
| 12 | Flygare Wallén, E., Ljunggren, G., Wahlström, L., Pettersson, D., Carlsson, A.C., Wändell, P. 2023 | The prevalence of self-harm and mental disorders among individuals with intellectual disabilities. Nordic Journal of Psychiatry*, 77*(7), 712–720. | Not a systematic review. It is a retrospective cohort study using administrative data. |
| 13 | Hedley, D., Hayward, S.M., Clarke, A., Uljarević, M., Stokes, M.A.2022 | Suicide and Autism: A Lifespan Perspective. In: Stancliffe, R.J., Wiese, M.Y., McCallion, P., McCarron, M. (eds) End of Life and People with Intellectual and Developmental Disability. Palgrave Macmillan, Cham. <https://doi.org/10.1007/978-3-030-98697-1_3> | Population not within scope, and not a systematic review. The paper provides a narrative overview of suicidality in autism, including risk/prevalence, role of intellectual disability, associated factors, assessment, and prevention, without focusing specifically on individuals with intellectual disability or using systematic methods |
| 14 | Hinze, E., Paynter, J., Dargue, N., Adams, D.2024 | The Presentation of Depression in Depressed Autistic Individuals: A Systematic Review, Rev J Autism Dev Disord. <https://doi.org/10.1007/s40489-024-00480-z> | Population not within scope of this review and report out of remit. It focuses on the presentation of depression in autistic individuals, with no prevalence data retrieved for the population with intellectual disability. |
| 15 | Hudson C., Chan J.2002 | Individuals with intellectual disability and mental illness: A literature review, Australian Journal of Social Issues, 37: 31-49. | Not a systematic review. It is a non-systematic literature review summarising existing research on the prevalence and treatment of mental illness in people with intellectual disability. |
| 16 | Lai, M.-C., Kassee, C., Besney, R., Bonato, S., Hull, L., Mandy, W., Szatmari, P., Ameis, S.H. 2019 | Prevalence of co-occurring mental health diagnoses in the autism population: a systematic review and meta-analysis. Lancet Psychiatry. Oct;6(10):819-829 | Population not within scope of this review and report out of remit. It focused on ASD population, and no prevalence data retrieved for the population with intellectual disability. ID considered as a moderator factor. |
| 17 | Madhavan, G.P.2018 | Psychotic symptoms in people with intellectual disability, *BMJ* 2018;360:k61 | Not a systematic review. It’s a letter to the Editor. |
| 18 | May, T., Pilkington, P.D., Younan, R., Williams, K.2021 | Overlap of autism spectrum disorder and borderline personality disorder: A systematic review and meta-analysis, Autism Res. Dec;14(12):2688-2710. | Population not within scope. It focuses on ASD and BPD, no prevalence data retrieved for the population with intellectual disability. |
| 19 | McGillicuddy N, Blane (1999) | Substance use in individuals with mental retardation. Addictive Behaviors. 1999; 24(6):869–878. | Not a systematic review. It is an expert overview of the literature. Specific search terms were provided, but no information was given regarding the databases consulted, the timeframe considered, or the inclusion/exclusion criteria applied. |
| 20 | Md Mahbub Hossain, et al. (2020) | Prevalence of comorbid psychiatric disorders among people with autism spectrum disorder: An umbrella review of systematic reviews and meta-analyses, Psychiatry Res. May;287:112922 | Not a systematic Review. It is an umbrella review on people with autism spectrum disorders. |
| 21 | Munir, Kerim Mm 2016 | The co-occurrence of mental disorders in children and adolescents with intellectual disability/intellectual developmental disorder, Curr Opin Psychiatry. Mar;29(2):95-102. | Not a systematic review. It is an expert, non-systematic overview of the literature regarding the comorbidity of mental disorders in children and adolescents with intellectual disability. |
| 22 | Shprintzen, R.J.2008 | Velo-cardio-facial syndrome: 30 Years of study, Dev Disabil Res Rev. 2008;14(1):3-10. | Not a systematic review. It is a non-systematic expert review of velo-cardio-facial syndrome. |
| 23 | Smiley E.2005 | Epidemiology of mental health problems in adults with learning disability: An update, Advances in Psychiatric Treatment.,11(3):214-222. doi:10.1192/apt.11.3.214 | Not a systematic review. It is an expert, non-systematic summary of the literature. |
| 24 | Tafolla, M., Lord, C.2024 | Longitudinal Analyses of Mental Health in Autistic Individuals: A Systematic Review, Brain Sci. 2024 Oct 18;14(10):1033 | Population not within scope of this review. It focuses on ASD population and no prevalence data have been retrieved for people with intellectual disabilities. |
| 25 | Totsika V, Liew A, Absoud M, Adnams C, Emerson E.,2022 | Mental health problems in children with intellectual disability. Lancet Child Adolesc Health. 2022 Jun;6(6):432-444. doi: 10.1016/S2352-4642(22)00067-0. Epub 2022 Apr 11. PMID: 35421380. | Not a systematic review. It is an umbrella review. |
| 26 | Kerim Munir, Ashok Roy, Afzal Javed Editors | Global e-Handbook of Intellectual Developmental Disorders. World Psychiatric Association. https://www.wpanet.org/books-produced-by-wpa | Not a systematic review. It is a book chapter identified through the World Psychiatric Association (WPA) website. |
| 27 | Galderisi S, Gorwood P, Gaebel W, Kurimay T, Dom G, Beezhold J, Wasserman D, Bailey S, Hanon C, Heinz A, Sartorius N, van der Gaag RJ, Vavrusova L, Wise J., 2018 | United Nations convention on the rights of persons with disabilities needs to be interpreted on the basis of scientific evidence regarding psychiatry. European Psychiatry. 2018 Sep;51:87-89. doi: 10.1016/j.eurpsy.2018.05.008. | Not a systematic review. It is a position paper. No prevalence data found. |
| 28 | Officer A, Shakespeare T., 2013 | The World Report on Disability and People With Intellectual Disabilities Journal of Policy and Practice in Intellectual Disabilities. 2013 Jun;10(2):79–81. doi: 10.1111/jppi.12031. | Not a systematic review. It is a report identified through the World Health Organization website |

#### Supplementary Table 7. Studies identified through database searching and included

| Alexander R & Cooray S., (2003) | Diagnosis of personality disorders in learning disability. Br J Psychiatry Suppl. 2003 Jan;44:S28-31. doi: 0.1192/bjp.182.44.s28. PMID: 12509306. |
| --- | --- |
| Aman H, Naeem F, Farooq S, Ayub M. (2016) | Prevalence of nonaffective psychosis in intellectually disabled clients: systematic review and meta-analysis. Psychiatr Genet. 2016 Aug;26(4):145-55. doi: 10.1097/YPG.0000000000000137. PMID: 27096221. |
| Bakken, T. L. and H. Martinsen (2013) | Adults with intellectual disabilities and mental illness in psychiatric inpatient units: Empirical studies of patient characteristics and psychiatric diagnoses from 1996 to 2011. International Journal of Developmental Disabilities, 59(3), 179–190. [https://doi.org/10.1179/2047387712Y.0000000006](https://psycnet.apa.org/doi/10.1179/2047387712Y.0000000006) |
| Curnow E, Rutherford M, Maciver D, Johnston L, Prior S, Boilson M, Shah P, Jenkins N, Meff T.  (2023) | Mental health in autistic adults: A rapid review of prevalence of psychiatric disorders and umbrella review of the effectiveness of interventions within a neurodiversity informed perspective. PLoS One. 2023 Jul 13;18(7):e0288275. doi: 10.1371/journal.pone.0288275. PMID: 37440543; PMCID: PMC10343158. |
| Daveney, J., Hassiotis, A., Katona, C., Matcham, F., & Sen, P. (2019). | Ascertainment and prevalence of post-traumatic stress disorder (PTSD) in people with intellectual Disabilities. Journal of Mental Health Research in Intellectual Disabilities, 12(3-4), 211–233. [https://doi.org/10.1080/19315864.2019.1637979](https://psycnet.apa.org/doi/10.1080/19315864.2019.1637979) |
| De Giorgi R, De Crescenzo F, D'Alò GL, Rizzo Pesci N, Di Franco V, Sandini C, Armando M. (2019) | Prevalence of Non-Affective Psychoses in Individuals with Autism Spectrum Disorders: A Systematic Review. J Clin Med. 2019 Aug 24;8(9):1304. doi: 10.3390/jcm8091304. PMID: 31450601; PMCID: PMC6780908. |
| Edwards G, Jones C, Pearson E, Royston R, Oliver C, Tarver J, Crawford H, Shelley L, Waite J. (2022) | Prevalence of anxiety symptomatology and diagnosis in syndromic intellectual disability: A systematic review and meta-analysis. Neurosci Biobehav Rev. 2022 Jul;138:104719. doi: 10.1016/j.neubiorev.2022.104719. Epub 2022 Jun 2. PMID: 35661754. |
| Glasson EJ, Buckley N, Chen W, Leonard H, Epstein A, Skoss R, Jacoby P, Blackmore AM, Bourke J, Downs J. (2020) | Systematic Review and Meta-analysis: Mental Health in Children With Neurogenetic Disorders Associated With Intellectual Disability. J Am Acad Child Adolesc Psychiatry. 2020 Sep;59(9):1036-1048. doi: 10.1016/j.jaac.2020.01.006. Epub 2020 Jan 13. PMID: 31945412. |
| Hollocks MJ, Lerh JW, Magiati I, Meiser-Stedman R, Brugha TS. (2019) | Anxiety and depression in adults with autism spectrum disorder: a systematic review and meta-analysis. Psychol Med. 2019 Mar;49(4):559-572. doi: 10.1017/S0033291718002283. Epub 2018 Sep 4. PMID: 30178724. |
| Huxley, A., Dalton, M., Tsui, Y. Y. Y., & Hayhurst, K. (2019) | Prevalence of alcohol, smoking and illicit drug use amongst people with Intellectual Disabilities: review. *Drugs: Education, Prevention and Policy*, *26*(5), 365-384. https://doi.org/10.1080/09687637.2018.1488949 |
| Maïano C, Coutu S, Tracey D, Bouchard S, Lepage G, Morin AJS, Moullec G.  (2018) | Prevalence of anxiety and depressive disorders among youth with intellectual disabilities: A systematic review and meta-analysis. J Affect Disord. 2018 Aug 15;236:230-242. doi: 10.1016/j.jad.2018.04.029. Epub 2018 Apr 6. PMID: 29751238. |
| Mazza MG, Rossetti A, Crespi G, Clerici M. (2020) | Prevalence of co-occurring psychiatric disorders in adults and adolescents with intellectual disability: A systematic review and meta-analysis. J Appl Res Intellect Disabil. 2020 Mar;33(2):126-138. doi: 10.1111/jar.12654. Epub 2019 Aug 20. PMID: 31430018. |
| Oeseburg B, Dijkstra GJ, Groothoff JW, Reijneveld SA, Jansen DE. (2011) | Prevalence of chronic health conditions in children with intellectual disability: a systematic literature review. Intellect Dev Disabil. 2011 Apr;49(2):59-85. doi: 10.1352/1934-9556-49.2.59. PMID: 21446871. |
| Rayner,K., Wood, H., Beail, N. and Nagra, M.K. (2015). | Intellectual disability, personality disorder and offending: a systematic review", Advances in Mental Health and Intellectual Disabilities, Vol. 9 No. 2, pp. 50-61. https://doi.org/10.1108/AMHID-04-2014-0007 |
| Royston R, Howlin P, Waite J, Oliver C. (2017) | Anxiety Disorders in Williams Syndrome Contrasted with Intellectual Disability and the General Population: A Systematic Review and Meta-Analysis. J Autism Dev Disord. 2017 Dec;47(12):3765-3777. doi: 10.1007/s10803-016-2909-z. PMID: 27696186; PMCID: PMC5676825. |
| Torr, J (2008) | Personality disorder and offending in people with learning disabilities", Advances in Mental Health and Learning Disabilities, Vol. 2 No. 1, pp. 4-10. https://doi.org/10.1108/17530180200800002 |
| van Duijvenbode N, VanDerNagel JEL. (2019) | A Systematic Review of Substance Use (Disorder) in Individuals with Mild to Borderline Intellectual Disability. Eur Addict Res. 2019;25(6):263-282. doi: 10.1159/000501679. Epub 2019 Jul 22. PMID: 31330514; PMCID: PMC6888885. |
| Varcin KJ, Herniman SE, Lin A, Chen Y, Perry Y, Pugh C, Chisholm K, Whitehouse AJO, Wood SJ (2022). | Occurrence of psychosis and bipolar disorder in adults with autism: A systematic review and meta-analysis. Neurosci Biobehav Rev. 2022 Mar;134:104543. doi: 10.1016/j.neubiorev.2022.104543. Epub 2022 Jan 19. PMID: 35063494. |
| Walton, C. and Kerr, M (2015). | Down syndrome: systematic review of the prevalence and nature of presentation of unipolar depression", Advances in Mental Health and Intellectual Disabilities, Vol. 9 No. 4, pp. 151-162. https://doi.org/10.1108/AMHID-11-2014-0037 |
| Walton C, Kerr M.  (2016) | Severe Intellectual Disability: Systematic Review of the Prevalence and Nature of Presentation of Unipolar Depression. J Appl Res Intellect Disabil. 2016 Sep;29(5):395-408. doi: 10.1111/jar.12203. Epub 2015 Jun 22. PMID: 26101049. |

#### Supplementary Table 8. Studies identified through manual searching and included

| **Authors, Year** | **Title, Journal, Vol, Iss** |
| --- | --- |
| Buckley N, Glasson EJ, Chen W, Epstein A, Leonard H, Skoss R, Jacoby P, Blackmore AM, Srinivasjois R, Bourke J, Sanders RJ, Downs J (2020) | Prevalence estimates of mental health problems in children and adolescents with intellectual disability: A systematic review and meta-analysis. Aust N Z J Psychiatry. 2020 Oct;54(10):970-984. doi: 10.1177/0004867420924101. Epub 2020 May 30. PMID: 32475125. |
| Jorge Lugo-Marín, María Magán-Maganto, Amado Rivero-Santana, Leticia Cuellar-Pompa, Montserrat Alviani, Cristina Jenaro-Rio, Emiliano Díez, Ricardo Canal-Bedia, (2019) | Prevalence of psychiatric disorders in adults with autism spectrum disorder: A systematic review and meta-analysis,  Research in Autism Spectrum Disorders, Volume 59, 2019, Pages 22-33, ISSN 1750-9467, https://doi.org/10.1016/j.rasd.2018.12.004. |
| Mevissen L., de Jongh A. (2010) | PTSD and its treatment in people with intellectual disabilities: A review of the literature. Clinical Psychology Review, Volume 30, Issue 3, 2010, Pages 308-316, ISSN 0272-7358, https://doi.org/10.1016/j.cpr.2009.12.005. |
| Reardon TC, Gray KM, Melvin GA (2015) | Anxiety disorders in children and adolescents with intellectual disability: Prevalence and assessment. Res Dev Disabil. 2015 Jan;36C:175-190. doi: 10.1016/j.ridd.2014.10.007. Epub 2014 Oct 21. PMID: 25462478. |
| van Steensel FJ, Bögels SM, Perrin S. (2011) | Anxiety disorders in children and adolescents with autistic spectrum disorders: a meta-analysis. Clin Child Fam Psychol Rev. 2011 Sep;14(3):302-17. doi: 10.1007/s10567-011-0097-0. PMID: 21735077; PMCID: PMC3162631. |
| Whitaker, S., & Read, S. (2006) | The Prevalence of Psychiatric Disorders among People with Intellectual Disabilities: An Analysis of the Literature. Journal of Applied Research in Intellectual Disabilities, 19(4), 330–345. https://doi.org/10.1111/j.1468-3148.2006.00293.x |

### Supplementary Table 9. PRISMA CHECKLIST

| **Section and Topic** | **Item #** | **Checklist item** | **Location where item is reported** |
| --- | --- | --- | --- |
| **TITLE** | | |  |
| Title | 1 | Identify the report as a systematic review. | N/A, it’s an umbrella review |
| **ABSTRACT** | | |  |
| Abstract | 2 | See the PRISMA 2020 for Abstracts checklist. | Yes |
| **INTRODUCTION** | | |  |
| Rationale | 3 | Describe the rationale for the review in the context of existing knowledge. | Background |
| Objectives | 4 | Provide an explicit statement of the objective(s) or question(s) the review addresses. | Background |
| **METHODS** | | |  |
| Eligibility criteria | 5 | Specify the inclusion and exclusion criteria for the review and how studies were grouped for the syntheses. | Methods, eligibility criteria |
| Information sources | 6 | Specify all databases, registers, websites, organisations, reference lists and other sources searched or consulted to identify studies. Specify the date when each source was last searched or consulted. | Methods, Literature sources |
| Search strategy | 7 | Present the full search strategies for all databases, registers and websites, including any filters and limits used. | Methods, literature sources, Supplementary Material Table 1 |
| Selection process | 8 | Specify the methods used to decide whether a study met the inclusion criteria of the review, including how many reviewers screened each record and each report retrieved, whether they worked independently, and if applicable, details of automation tools used in the process. | Methods, Eligibility criteria, Procedure, Fig.1, |
| Data collection process | 9 | Specify the methods used to collect data from reports, including how many reviewers collected data from each report, whether they worked independently, any processes for obtaining or confirming data from study investigators, and if applicable, details of automation tools used in the process. | Methods, Procedure, quality appraisal |
| Data items | 10a | List and define all outcomes for which data were sought. Specify whether all results that were compatible with each outcome domain in each study were sought (e.g. for all measures, time points, analyses), and if not, the methods used to decide which results to collect. | Methods, procedure |
|  | 10b | List and define all other variables for which data were sought (e.g. participant and intervention characteristics, funding sources). Describe any assumptions made about any missing or unclear information. | Methods, procedure |
| Study risk of bias assessment | 11 | Specify the methods used to assess risk of bias in the included studies, including details of the tool(s) used, how many reviewers assessed each study and whether they worked independently, and if applicable, details of automation tools used in the process. | Methods, SM Table 2-4. |
| Effect measures | 12 | Specify for each outcome the effect measure(s) (e.g. risk ratio, mean difference) used in the synthesis or presentation of results. | Methods |
| Synthesis methods | 13a | Describe the processes used to decide which studies were eligible for each synthesis (e.g. tabulating the study intervention characteristics and comparing against the planned groups for each synthesis (item #5)). | Methods |
|  | 13b | Describe any methods required to prepare the data for presentation or synthesis, such as handling of missing summary statistics, or data conversions. | Methods |
|  | 13c | Describe any methods used to tabulate or visually display results of individual studies and syntheses. | Methods |
|  | 13d | Describe any methods used to synthesize results and provide a rationale for the choice(s). If meta-analysis was performed, describe the model(s), method(s) to identify the presence and extent of statistical heterogeneity, and software package(s) used. | Methods |
|  | 13e | Describe any methods used to explore possible causes of heterogeneity among study results (e.g. subgroup analysis, meta-regression). | NA |
|  | 13f | Describe any sensitivity analyses conducted to assess robustness of the synthesized results. | N/A |
| Reporting bias assessment | 14 | Describe any methods used to assess risk of bias due to missing results in a synthesis (arising from reporting biases). | Methods, SM Table 2-4. |
| Certainty assessment | 15 | Describe any methods used to assess certainty (or confidence) in the body of evidence for an outcome. | Methods |
| **RESULTS** | | |  |
| Study selection | 16a | Describe the results of the search and selection process, from the number of records identified in the search to the number of studies included in the review, ideally using a flow diagram. | Results, Fig.1 |
|  | 16b | Cite studies that might appear to meet the inclusion criteria, but which were excluded, and explain why they were excluded. | Results, Supplementary Materials Table 5-6 |
| Study characteristics | 17 | Cite each included study and present its characteristics. | Results, Table 1-2, SM Table 2-4, 7-8 |
| Risk of bias in studies | 18 | Present assessments of risk of bias for each included study. | NA. Prevalence rates discussed in comparison to the general population. |
| Results of individual studies | 19 | For all outcomes, present, for each study: (a) summary statistics for each group (where appropriate) and (b) an effect estimate and its precision (e.g. confidence/credible interval), ideally using structured tables or plots. | Results, Table 1-3. |
| Results of syntheses | 20a | For each synthesis, briefly summarise the characteristics and risk of bias among contributing studies. | Results, Table 1, Discussion |
|  | 20b | Present results of all statistical syntheses conducted. If meta-analysis was done, present for each the summary estimate and its precision (e.g. confidence/credible interval) and measures of statistical heterogeneity. If comparing groups, describe the direction of the effect. | NA |
|  | 20c | Present results of all investigations of possible causes of heterogeneity among study results. | Results, Discussion, SM Table 2-4 |
|  | 20d | Present results of all sensitivity analyses conducted to assess the robustness of the synthesized results. | NA |
| Reporting biases | 21 | Present assessments of risk of bias due to missing results (arising from reporting biases) for each synthesis assessed. | NA. |
| Certainty of evidence | 22 | Present assessments of certainty (or confidence) in the body of evidence for each outcome assessed. | Results, Table 2 |
| **DISCUSSION** | | |  |
| Discussion | 23a | Provide a general interpretation of the results in the context of other evidence. | Discussion |
|  | 23b | Discuss any limitations of the evidence included in the review. | Discussion |
|  | 23c | Discuss any limitations of the review processes used. | Strengths and limitations |
|  | 23d | Discuss implications of the results for practice, policy, and future research. | Discussion, Clinical and Policy Implications |
| **OTHER INFORMATION** | | |  |
| Registration and protocol | 24a | Provide registration information for the review, including register name and registration number, or state that the review was not registered. | Abstract, Methods |
|  | 24b | Indicate where the review protocol can be accessed, or state that a protocol was not prepared. | Methods |
|  | 24c | Describe and explain any amendments to information provided at registration or in the protocol. | SM Table 1 |
| Support | 25 | Describe sources of financial or non-financial support for the review, and the role of the funders or sponsors in the review. | Funding |
| Competing interests | 26 | Declare any competing interests of review authors. | Declaration of Interest |
| Availability of data, code and other materials | 27 | Report which of the following are publicly available and where they can be found: template data collection forms; data extracted from included studies; data used for all analyses; analytic code; any other materials used in the review. | Data Availability |

*From:* Page MJ, McKenzie JE, Bossuyt PM, Boutron I, Hoffmann TC, Mulrow CD, et al. The PRISMA 2020 statement: an updated guideline for reporting systematic reviews. BMJ 2021;372:n71. doi: 10.1136/bmj.n71

### Supplementary Table 10. PRISMA 2020 for Abstracts Checklist

| **Section and Topic** | **Item #** | **Checklist item** | **Reported (Yes/No)** |
| --- | --- | --- | --- |
| **TITLE** | | |  |
| Title | 1 | Identify the report as a systematic review. | N/A |
| **BACKGROUND** | | |  |
| Objectives | 2 | Provide an explicit statement of the main objective(s) or question(s) the review addresses. | Yes |
| **METHODS** | | |  |
| Eligibility criteria | 3 | Specify the inclusion and exclusion criteria for the review. | Yes |
| Information sources | 4 | Specify the information sources (e.g. databases, registers) used to identify studies and the date when each was last searched. | Yes |
| Risk of bias | 5 | Specify the methods used to assess risk of bias in the included studies. | Yes |
| Synthesis of results | 6 | Specify the methods used to present and synthesise results. | Yes |
| **RESULTS** | | |  |
| Included studies | 7 | Give the total number of included studies and participants and summarise relevant characteristics of studies. | Yes |
| Synthesis of results | 8 | Present results for main outcomes, preferably indicating the number of included studies and participants for each. If meta-analysis was done, report the summary estimate and confidence/credible interval. If comparing groups, indicate the direction of the effect (i.e. which group is favoured). | Yes |
| **DISCUSSION** | | |  |
| Limitations of evidence | 9 | Provide a brief summary of the limitations of the evidence included in the review (e.g. study risk of bias, inconsistency and imprecision). | Yes |
| Interpretation | 10 | Provide a general interpretation of the results and important implications. | Yes |
| **OTHER** | | |  |
| Funding | 11 | Specify the primary source of funding for the review. | Yes |
| Registration | 12 | Provide the register name and registration number. | Yes |

*From:* Page MJ, McKenzie JE, Bossuyt PM, Boutron I, Hoffmann TC, Mulrow CD, et al. The PRISMA 2020 statement: an updated guideline for reporting systematic reviews. BMJ 2021;372:n71. doi: 10.1136/bmj.n71. This work is licensed under CC BY 4.0. To view a copy of this license, visit <https://creativecommons.org/licenses/by/4.0/>
